# A Genome-wide Genetic Data Resource for the 45 and Up Study

**DOI:** 10.64898/2026.09.20.26363524

**Authors:** Hamzeh Mesrian Tanha, David Goldsbury, Richard Parker, Natalie Garden, Erika de Guzman, Greer Dawson, Kerrin Bleicher, Alison Cowle, Martin McNamara, Anne E Cust, Nicholas G Martin, Julia Steinberg

## Abstract

Large population-based cohorts enable investigation of disease risk and precision prevention, but often lack genome-wide genetic data. We generated and validated new genomic data within the Australian 45 and Up Study, one of the largest Southern Hemisphere cohorts (recruited 2005-2009) with extensive longitudinal and linked health data.

In 2022-2023, 30,541 participants were invited (random sub-cohort and all individuals with history of four most common invasive cancers: breast, prostate, melanoma, colorectal), with in-depth mapping of participants’ characteristics for people who consented to provide a DNA sample and those who did not. Low-coverage whole-genome sequencing data (0.4-6.3×) followed by imputation yielded ∼79 million variants for n=7,408 individuals; data for n=6,827 passed stringent quality control. Genotype concordance was high between duplicates (n=85) and with dense genotyping arrays (n=188). Most unrelated participants with high-quality data had inferred European genetic ancestry (n=6,631, 98%), with n=141 (2%) of inferred non-European or admixed ancestry. As proof-of-principle integration of new genomic data with extensive existing linked health records, polygenic risk scores (PGS) for four most common cancers showed predictive performance broadly consistent with previous studies, including association between PGS and earlier age at prostate cancer diagnosis, and no meaningful differences in cancer stage at diagnosis by PGS.

This new resource has substantially enhanced the 45 and Up Study, supporting investigations of disease aetiology, risk stratification, and precision health in Australia and globally.

## 1. Introduction

Characterising genetic variation and its contribution to health and disease (alongside environmental, behavioural and sociodemographic determinants) is a central goal of population health research^1, 2^. Longitudinal cohorts that couple genomic profiling with deep phenotyping and record linkage have transformed our ability to dissect disease aetiology and improve risk prediction across populations^3, 4^. However, many well-characterised cohorts still lack genome-wide genetic data, limiting genetic and gene–environment analyses. The Sax Institute’s 45 and Up Study is one of the largest cohorts in the Southern Hemisphere, comprising 267,357 residents of NSW, Australia, who were recruited in 2005–2009 and aged ≥45 years^5^. This cohort includes in-depth baseline and follow-up information on health, lifestyle and socioeconomic factors, with linkage to extensive population-wide records including deaths, cancers, hospital admissions, general practice and specialist visits, medical procedures and pharmaceutical dispensing through Australia’s universal health insurance scheme (Medicare)^5^. Despite this breadth of longitudinal data, genome-wide genetic data have only been generated for a small, highly selected subset of healthy older individuals^6^, creating a key gap for a resource that has already been used in >500 research publications and informed work by >30 policy agencies^7^. Generation of genome-wide information in this cohort can help strengthen Australian and international efforts to elucidate the genetic architecture of complex disease, refine disease risk predictions, and inform targeted prevention and intervention strategies to improve health outcomes.

Advances in technology have made low-coverage whole-genome sequencing (lcWGS) a practical, cost-efficient alternative to genotyping arrays for large-scale genetic studies^8, 9, 10^. At shallow depth (typically <1×), lcWGS combined with genotype imputation from large reference panels has been shown to recover genome-wide variation (including less-common variants) with high accuracy, often at similar or lower per-sample cost than dense genotyping arrays, with lcWGS at ∼0.5× coverage able to generate data sufficiently accurate for downstream analyses without requiring aggressive filtering^12^. Across diverse populations, lcWGS-based imputation at 0.5× coverage has been reported to perform similarly to or even outperform array-based imputation^11^, particularly for common variants, while reducing array ascertainment bias arising from designs optimised for European populations^12^.

In this context, we used lcWGS to generate genome-wide genetic data for >7,400 participants of the 45 and Up Study, creating a new valuable resource for population and disease genetics research. Participants were recruited through a combination of random sampling and targeted invitation of individuals with four common cancers, supporting both broad future research and cancer-focused analyses as part of the Australian Cancer Risk Study (cancerrisk.au; a transdisciplinary program evaluating the potential of genomics-informed, risk-based cancer screening). Here, we describe this new genetic dataset and its integration with rich longitudinal phenotypic and linked health records. Notably, by embedding the DNA sample collection within an existing cohort, we provide an in-depth examination of participants’ characteristics for people who provided a DNA sample and those who did not. We further demonstrate genetic data quality, including through validation via genotype array data, and showcase utility through linkage-enabled, proof-of-concept evaluations of polygenic risk scores for four major cancers.

## 2. Results

### 2.1 Participants’ characteristics

Between 2022 and 2023, we invited 30,541 participants of the 45 and Up Study to provide a saliva sample for DNA analysis. Invitations included a randomly selected sub-cohort (n=9,986) and all participants diagnosed with invasive prostate, breast, melanoma, or colorectal cancer who were alive as of October 2021 (Fig. 1). We tested and refined an invitation process in a pilot set of 2,400 participants, evaluating consent rates after initial invitations and up to 2 reminders, and different consent models (see Supplementary Information). In particular, we found highly similar consent rates (<2% difference) for “fine-grained” consent (i.e. separate consent to this study and future health/medical research) and “broad” consent (i.e. consent to this study and future health/medical research), thus proceeded with the combined “broad” consent model. In total, 8,311 individuals (27% of those invited) consented to provide a DNA sample, with different consent rates by invitation mode (39% for email invitation, 1 email reminder, 1 postal reminder; 17% for postal invitation, 1 postal reminder). Consent rates were slightly higher for participants with history of breast, prostate cancer or melanoma (28-29%) than for participants with history of colorectal cancer (24%) or randomly selected participants with no history of the four major cancers (25%; see Supplementary Information for detailed consent rates).

**Figure 1.**
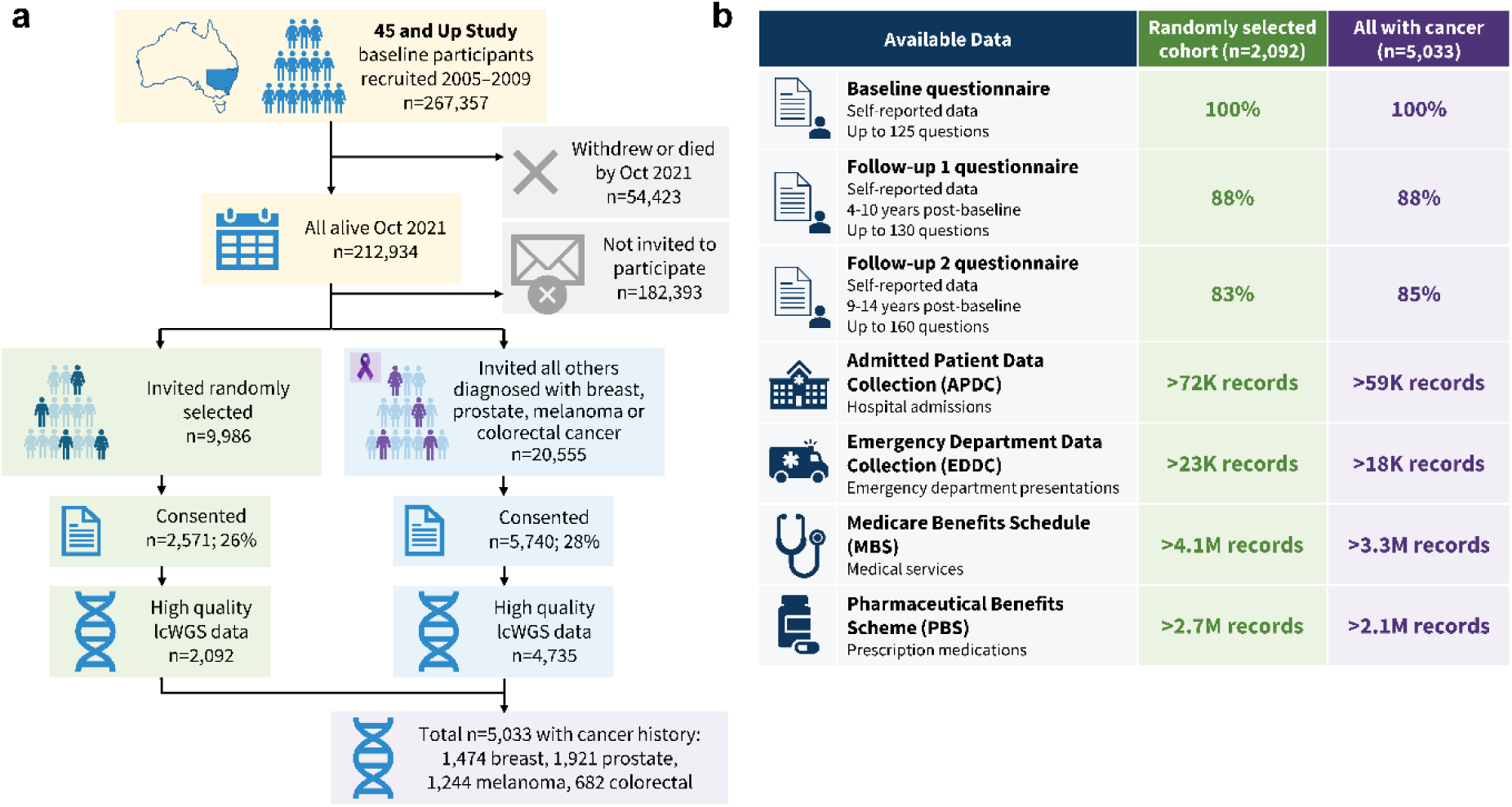
Overview of participant recruitment, genomic data generation, and linked health data in the 45 and Up Study. **(a)** Participant recruitment and generation of the genomic dataset. Of the 212,934 participants alive in October 2021, 30,541 were invited to provide saliva samples for genomic analysis, including a randomly selected sub-cohort (n=9,986) and all remaining participants diagnosed with breast, prostate, colorectal, or melanoma cancer (n=20,555). Following consent, low-coverage whole-genome sequencing (lcWGS), genotype imputation, and stringent quality checks, the final high-quality genomic dataset comprised 6,827 participants, including 2,092 from the randomly selected sub-cohort and 5,033 participants with a history of one or more of the four cancers (1,474 breast, 1,921 prostate, 1,244 melanoma, and 682 colorectal cancer). **(b)** Summary of questionnaire and linked administrative health data available for participants with high-quality lcWGS data in the randomly selected cohort (n=2,092) and all participants with cancer (n=5,033). Note, participants with cancer may also be included in the randomly selected sub-cohort. Data sources include baseline and follow-up questionnaires, the Admitted Patient Data Collection, Emergency Department Data Collection, Medicare Benefits Schedule, and Pharmaceutical Benefits Scheme. Questionnaire values indicate participant availability, whereas other values represent the total number of linked records. Definitions of questionnaire items and response categories were based on the 45 and Up Study questionnaires and data dictionaries, available from the Sax Institute website^13^.

Using in-depth previous questionnaire data for invited and consented individuals (Table 1; Supplementary Table 2), we mapped sociodemographic and health-related characteristics associated with providing consent (Fig. 2), with simultaneous adjustment for the wide range of characteristics included in the analysis. Providing consent was significantly (*p*<0.001) associated with several key health factors, including self-reported excellent health at baseline (adjusted odds ratio (aOR)=1.77 (95% confidence interval (CI):1.56-2.00) versus fair/poor health) and a non-linear association with age at sample collection (e.g. aOR=1.30 (95%CI: 1.15-1.45) for 70-74 versus 60-64 years, but aOR=0.69 (95%CI:0.61-0.79) for 85-89 versus 60-64 years). There was no association with gender (*p*>0.1). Considering socioeconomic advantage, providing consent was significantly associated with both education (e.g. aOR=1.75 (95%CI: 1.62-1.89) for university versus did not finish high school) and higher household income (aOR=1.21 (95%CI:1.11-1.32) for >$70,000 versus <$30,000), but not with area-based socioeconomic disadvantage or private health insurance status (both *p*>0.10), and was not lower for more remote areas of residence (Fig. 2). People with darker skin colour (self-reported dark olive/brown/black aOR=0.58 (95%CI: 0.48-0.70) versus very fair) and birth overseas (aOR=0.83 (95%CI:0.77-0.89) versus Australia) were also less likely to provide consent.

**Table 1.** Demographic and health-related characteristics of 45 and Up Study participants invited, those who consented, those with high-quality genomic data, and unrelated individuals with high-quality data and inferred European genetic ancestry.

|  | <u>Invited</u> |  | <u>Consented</u> |  | <u>High quality genomic data</u> |  | <u>High quality data, unrelated, European ancestry</u> |  |
| --- | --- | --- | --- | --- | --- | --- | --- | --- |
| **Characteristic** | **n** | **Column %** | **n** | **% of invited** | **n** | **% of invited** | **n** | **% of invited** |
| **All participants** | 30,541 | 100% | 8,311 | 27% | 6,827 | 22% | 6,631 | 22% |
| Randomly selected cohort | 9,986 | 33% | 2,571 | 26% | 2,092 | 21% | 2,011 | 20% |
| Breast cancer group* | 5,628 | 18% | 1,629 | 29% | 1,371 | 24% | 1,335 | 24% |
| Colorectal cancer group* | 2,967 | 10% | 713 | 24% | 582 | 20% | 564 | 19% |
| Melanoma group* | 4,556 | 15% | 1,282 | 28% | 1,072 | 24% | 1,048 | 23% |
| Prostate cancer group* | 7,404 | 24% | 2,116 | 29% | 1,710 | 23% | 1,673 | 23% |
| **Gender** |  |  |  |  |  |  |  |  |
| Female | 14,839 | 49% | 4,004 | 27% | 3,364 | 23% | 3,256 | 22% |
| Male | 15,702 | 51% | 4,307 | 27% | 3,463 | 22% | 3,375 | 21% |
| **Age at baseline (years)** |  |  |  |  |  |  |  |  |
| 45-54 | 7,894 | 26% | 2,468 | 31% | 2,040 | 26% | 1,968 | 25% |
| 55-64 | 11,941 | 39% | 3,716 | 31% | 3,112 | 26% | 3,026 | 25% |
| 65-74 | 8,507 | 28% | 1,873 | 22% | 1,485 | 17% | 1,452 | 17% |
| 75+ | 2,199 | 7% | 254 | 12% | 190 | 9% | 185 | 8% |
| **Age at end of DNA sample collection (years)** |  |  |  |  |  |  |  |  |
| 55-64 | 2,556 | 8% | 705 | 28% | 581 | 23% | 552 | 22% |
| 65-74 | 10,249 | 34% | 3,344 | 33% | 2,780 | 27% | 2,698 | 26% |
| 75-84 | 11,584 | 38% | 3,289 | 28% | 2,711 | 23% | 2,643 | 23% |
| 85+ | 6,152 | 20% | 973 | 16% | 755 | 12% | 738 | 12% |
| **Highest educational qualification** |  |  |  |  |  |  |  |  |
| University degree or higher | 7,869 | 26% | 2,897 | 37% | 2,378 | 30% | 2,296 | 29% |
| Certificate/Diploma | 6,676 | 22% | 1,984 | 30% | 1,661 | 25% | 1,619 | 24% |
| Trade/Apprenticeship | 3,481 | 11% | 806 | 23% | 660 | 19% | 643 | 18% |
| High school | 2,848 | 9% | 715 | 25% | 578 | 20% | 565 | 20% |
| Did not complete high school | 9,307 | 30% | 1,858 | 20% | 1,511 | 16% | 1,469 | 16% |
| Unknown | 360 | 1% | 51 | 14% | 39 | 11% | 39 | 11% |
| **Socioeconomic index of place of residence** |  |  |  |  |  |  |  |  |
| Most disadvantaged quintile | 5,398 | 18% | 1,259 | 23% | 1,007 | 19% | 979 | 18% |
| Quintile 2 | 6,062 | 20% | 1,516 | 25% | 1,244 | 21% | 1,215 | 20% |
| Quintile 3 | 5,695 | 19% | 1,551 | 27% | 1,277 | 22% | 1,242 | 22% |
| Quintile 4 | 5,529 | 18% | 1,598 | 29% | 1,338 | 24% | 1,294 | 23% |
| Least disadvantaged quintile | 7,042 | 23% | 2,130 | 30% | 1,740 | 25% | 1,685 | 24% |
| Unknown | 815 | 3% | 257 | 32% | 221 | 27% | 216 | 27% |
| **Accessibility/remoteness of place of residence** |  |  |  |  |  |  |  |  |
| Major cities | 16,004 | 52% | 4,324 | 27% | 3,566 | 22% | 3,433 | 21% |
| Inner regional | 10,692 | 35% | 2,913 | 27% | 2,382 | 22% | 2,336 | 22% |
| Outer regional / Remote / Very remote | 3,460 | 11% | 945 | 27% | 771 | 22% | 759 | 22% |
| Unknown | 385 | 1% | 129 | 34% | 108 | 28% | 103 | 27% |
| **Country of birth** |  |  |  |  |  |  |  |  |
| Australia | 23,859 | 78% | 6,682 | 28% | 5,547 | 23% | 5,433 | 23% |
| Other | 6,478 | 21% | 1,576 | 24% | 1,240 | 19% | 1,158 | 18% |
| Unknown | 204 | 1% | 53 | 26% | 40 | 20% | 40 | 20% |
| **Self-reported health** |  |  |  |  |  |  |  |  |
| Excellent | 4,886 | 16% | 1,745 | 36% | 1,453 | 30% | 1,421 | 29% |
| Very good | 12,247 | 40% | 3,600 | 29% | 3,005 | 25% | 2,908 | 24% |
| Good | 9,776 | 32% | 2,308 | 24% | 1,861 | 19% | 1,809 | 19% |
| Fair/Poor | 2,782 | 9% | 497 | 18% | 383 | 14% | 373 | 13% |
| Unknown | 850 | 3% | 161 | 19% | 125 | 15% | 120 | 14% |
| **Personal history of diseases** |  |  |  |  |  |  |  |  |
| Heart disease/Stroke | 3,298 | 11% | 725 | 22% | 580 | 18% | 564 | 17% |
| High blood pressure | 10,766 | 35% | 2,494 | 23% | 2,048 | 19% | 2,001 | 19% |
| Diabetes | 2,207 | 7% | 421 | 19% | 334 | 15% | 322 | 15% |
| Depression/anxiety | 4,411 | 14% | 1,233 | 28% | 1,014 | 23% | 981 | 22% |
| Blood clots/thrombosis | 1,291 | 4% | 338 | 26% | 278 | 22% | 273 | 21% |
| **Family history of cancer** |  |  |  |  |  |  |  |  |
| Breast cancer | 3,855 | 13% | 1,126 | 29% | 924 | 24% | 886 | 23% |
| Colorectal cancer | 4,576 | 15% | 1,215 | 27% | 1,001 | 22% | 976 | 21% |
| Melanoma | 3,213 | 11% | 1,011 | 31% | 858 | 27% | 839 | 26% |
| Prostate cancer | 3,905 | 13% | 1,130 | 29% | 944 | 24% | 929 | 24% |
| Lung cancer | 3,148 | 10% | 867 | 28% | 710 | 23% | 694 | 22% |
| Ovarian cancer | 1,079 | 4% | 288 | 27% | 227 | 21% | 221 | 20% |
| Any of the above | 14,689 | 48% | 4,174 | 28% | 3,430 | 23% | 3,338 | 23% |

Notes: Some smaller cells and the cells around them have been perturbed to preserve privacy (e.g. where the change from all with high quality genomic data to those with European ancestry was <5). Data sources and questionnaire items underlying all characteristics are shown in Supplementary Table 1. Additional characteristics are listed in Supplementary Table 2. \* not including participants in randomly selected cohort; each individual assigned to group based on the first cancer diagnosis.

**Figure 2.**
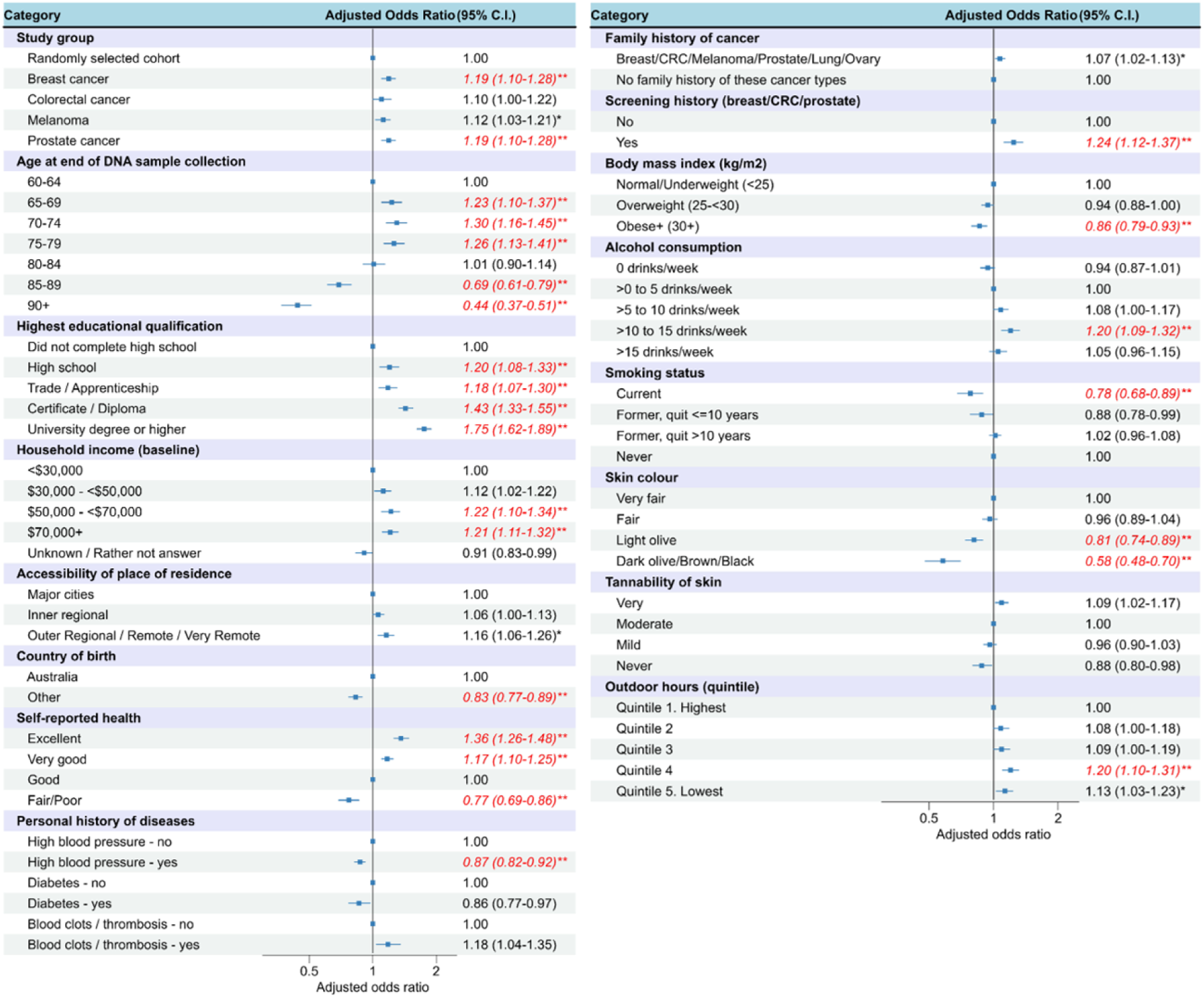
Participants’ characteristics associated with consent to provide DNA sample, among n=30,541 invited participants (n=G,G86 randomly selected, and all participants alive with a history of breast, prostate, melanoma, or colorectal cancer). Regression analysis with simultaneous adjustment for all characteristics shown in the figure, based on step-wise forward selection (p<0.1). Characteristics not retained by step-wise forward selection (p>0.1) were health insurance status, area-level index of socioeconomic disadvantage, gender, and personal history of heart disease/stroke, personal history of depression/anxiety. ** p<0.001; * p<0.01; CRC: colorectal cancer.

The Supplementary Information includes further results for characteristics associated with genomic data being included in the high-quality dataset passing all quality checks (as described below) among all individuals who consented.

### 2.2 Overview of genomic data

In total, 7,941 participants (96% of those who consented) returned a DNA sample (Supplementary Fig. 1). Of those, lcWGS data with minimum 0.4× average coverage and passing initial quality checks (QC; see Methods) were successfully generated for 7,408 individuals (93% of those who returned a DNA sample), with added technical duplicates for 100 participants (n=91: duplicate independently sequenced and imputed; n=9: duplicate independently imputed using same sequencing data). Genotype imputation (combined 1000 Genomes^14^, HGDP^15^ and gnomAD^16^ reference panel) yielded ∼79.3 million variants reported in VCF files.

We further evaluated genotype quality for these data (Fig. 3; Supplementary Figs. 2–3). Raw sequencing coverage was strongly positively associated with the number of reference-panel variant sites covered by ≥1 read (*r*=0.62, *p*<2.2×10^−16^; Fig. 3a).

**Figure 3.**
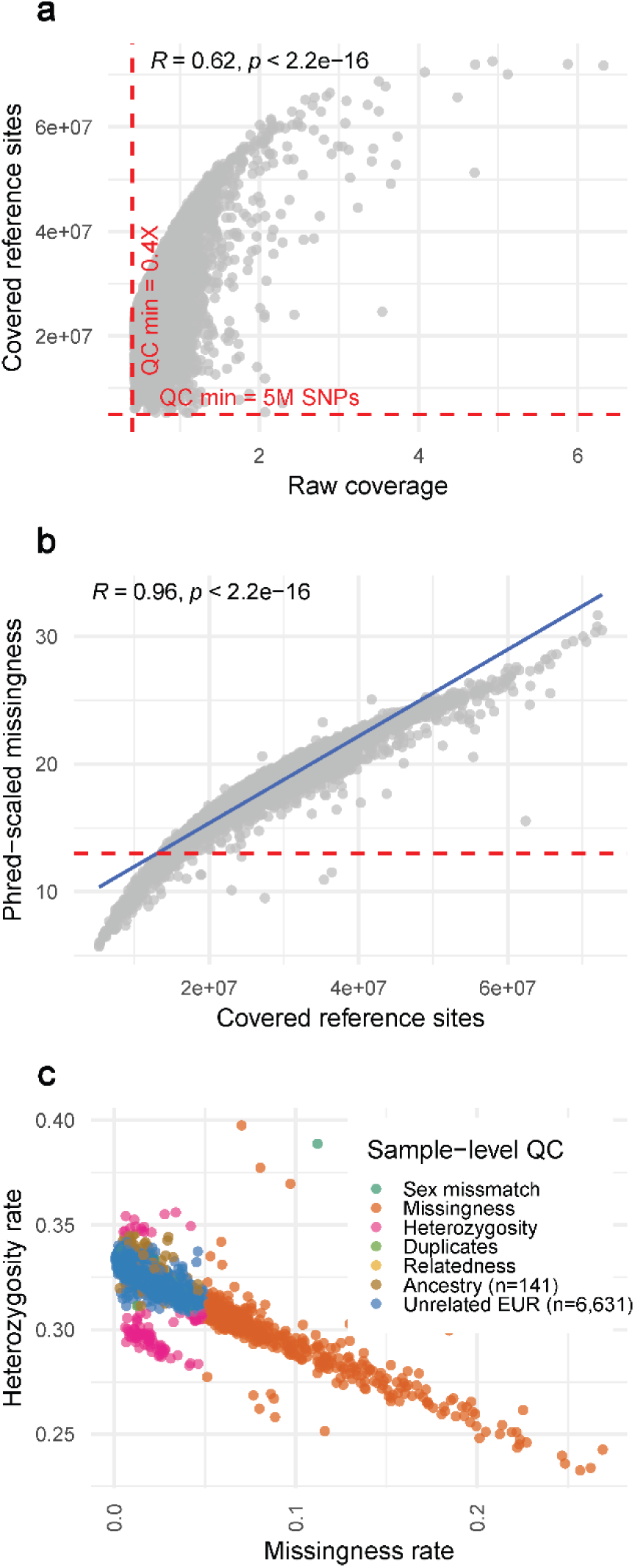
Individual-level quality control of newly generated low-coverage whole-genome sequencing data. **(a)** Relationship between raw sequencing coverage and the number of reference-panel variant sites covered by ≥1 sequencing read. All samples satisfied minimum thresholds of >0.4x raw coverage and >5 million reference-panel sites covered by ≥1 read (per initial QC within data generation). **(b)** Association between the number of covered reference-panel variant sites and phred-scaled genotype missingness (“missing” genotypes defined as GP<0.9). The dashed line indicates a missingness threshold of ≤5% of variants with GP<0.9 for an individual (phred ≈ 13), which corresponds to ∼13.7 million reference-panel variant sites covered by ≥1 read. Raw coverage showed consistent but weaker association with missingness (Supplementary Fig. 2a). Panels **a–b** are based on all lcWGS data, including 7,408 unique individuals and 100 duplicate samples. **(c)** Sample-level quality control steps based on genotype missingness and heterozygosity rates. Samples failing QC due to sex mismatch (genetic vs reported sex), high missingness (>5% per sample), extreme heterozygosity (outside mean ±3 SD), duplicates, or relatedness (retaining one individual from each duplicate or 1st/2nd-degree relative pair) were excluded. After quality control and removal of related individuals, 6,631 unrelated individuals of European genetic ancestry and 141 unrelated individuals of predominantly non-European or admixed ancestry were identified. European genetic ancestry was defined as PC1–PC4 values within ±3 SD of the corresponding means in 1000 Genomes European reference individuals.

To establish data quality and for further downstream analyses, we retained samples with high-confidence imputation (genotype probability GP≥0.9) for ≥95% of post-imputation variants across a combined set of HapMap3 and leading cancer polygenic risk scores (PGS; here and throughout, 7 PGS for breast, prostate, melanoma and colorectal cancers, with 68-451 variants each; see Methods). This corresponded to minimum ∼13.7 million reference panel variants covered by ≥1 read (Fig. 3b), and a minimum raw coverage of ∼0.426× (Supplementary Fig. 2a). We further removed 8% of samples due to sex discordance, elevated missingness, and extreme heterozygosity (see Methods). Relatedness estimates based on KING-robust identified known duplicate samples and likely parent–child pairs and, separately, full sibling pairs (Supplementary Fig. 3); these classifications were supported by reported years of birth in existing 45 and Up Study data (parent–child pairs typically differing by >20 years, siblings by <10 years). After exclusion of duplicates, we retained a high-quality genetic dataset of 6,827 individuals, including 6,772 unrelated participants (Table 1, Fig. 3c).

Among the 6,772 unrelated participants with high-quality data, based on principal component analysis (PCA), 6,631 (98%) were inferred to be of European genetic ancestry and 141 (2%) to be of non-European or substantially admixed ancestry (Fig. 3c, Fig. 4a–b). While self-reported ancestry information for participants is somewhat limited (questionnaire with tick boxes for a small number of ancestries common in Australia, including a broad “Australian” category selected by the majority of all participants, and a broad “other” category), the inferred genetic ancestry was also broadly consistent with self-reported ancestry. Among the 6,631 participants inferred to be of European genetic ancestry, only 0.6% self-reported at least one specific non-European and non-Australian ancestry category in baseline questionnaires (Chinese, Filipino, Indian, Lebanese, Maltese or Vietnamese) and 7.4% selected “other” ancestry (noting this could include less common European ancestries). By contrast, for the 141 participants with inferred non-European or admixed genetic ancestry, relevant proportions were 29% and 61%, respectively.

**Figure 4.**
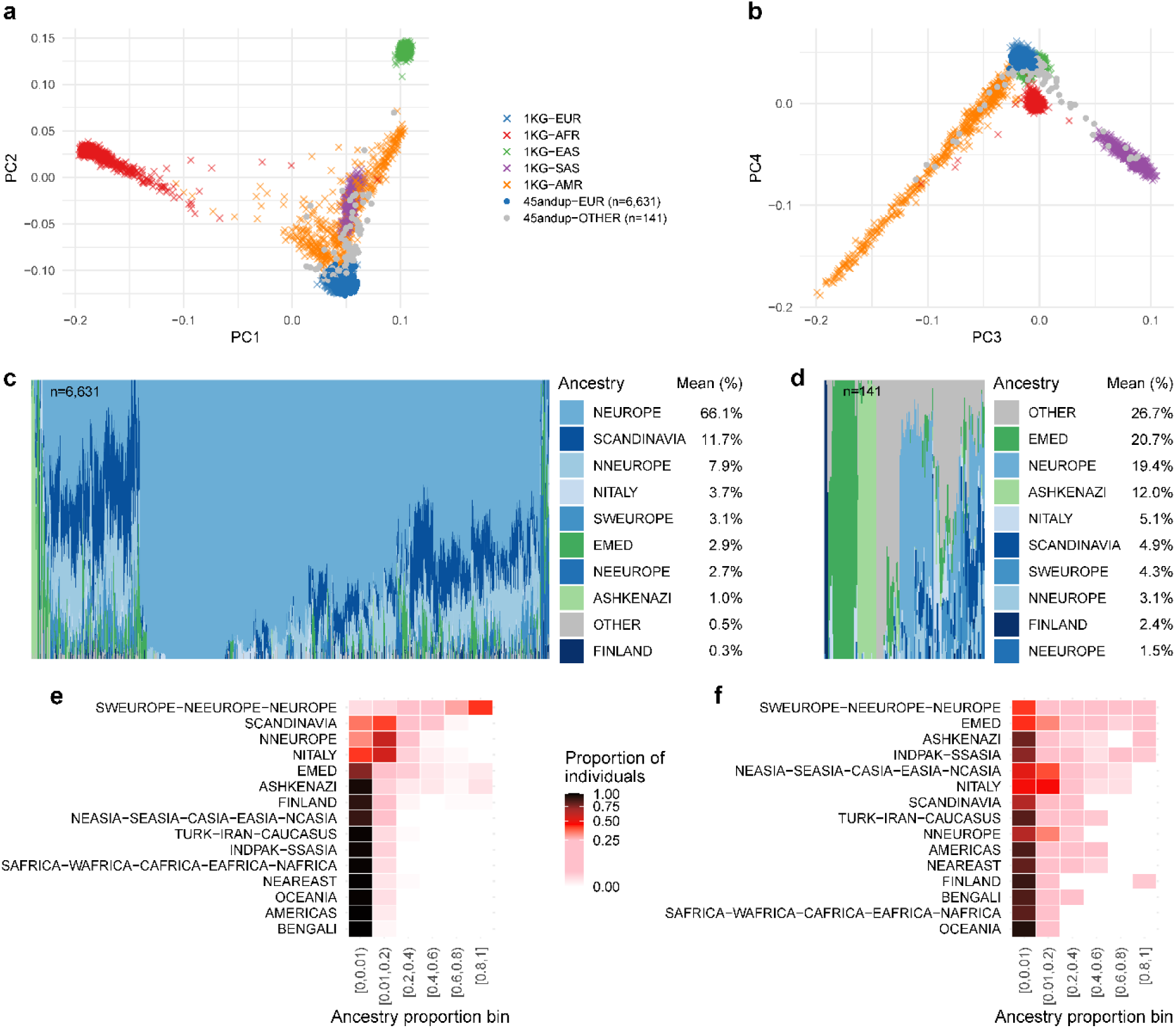
Genetic ancestry inference and population structure of the 45 and Up Study genomic dataset. Principal component analysis (PCA) and supervised ancestry deconvolution (Gencove proprietary model with custom reference panel) were used to characterise the genetic ancestry of study participants. **(a–b)** Projection of 45 and Up Study participants (unrelated individuals with high-quality genomic data, n = 6,772) onto principal components derived from the 1000 Genomes reference populations (**a**, PC1 vs PC2; **b**, PC3 vs PC4). Participants predominantly cluster with individuals of European ancestry (n = 6,631; within 3sd of mean), with a smaller subset outside the cluster, suggesting non-European or admixed ancestry (n = 141). **(c)** Complementary genetic ancestry deconvolution analysis of individuals classified as European-ancestry (n = 6,631) based on PCA. **(d)** Genetic ancestry deconvolution of individuals classified as non-European or admixed ancestry (n = 141) in PCA analysis. Figure shows same ancestry components as in panel **c**, ordered according to their mean ancestry contribution, to facilitate direct comparison between groups. **(e–f)** Distribution of ancestry proportions for individuals of inferred European ancestry (**e**; n = 6,631) and individuals of inferred non-European or admixed ancestry (**f**; n = 141), based on Gencove algorithm estimates. Heatmap colours indicate the proportion of individuals within each ancestry component whose inferred ancestry contribution falls within a given ancestry proportion bin. The ancestry groups are ordered according to their mean contribution.

Moreover, the ancestry assignment was generally consistent with results from a complementary ancestry deconvolution using a Gencove proprietary algorithm; for example, the latter indicated that individuals assigned European ancestry in PCA on average comprised 66% Northern/Central European (British, Irish, German) and 11% Scandinavian (Icelandic, Norwegian, Swedish) ancestry (Fig. 4c), with 42% of individuals having >80% combined Southwestern European, Northeastern European, and Northern/Central European ancestry (Fig 4e). By contrast, for the 141 non-European or admixed individuals based on PCA, ancestry contributions inferred by the deconvolution analysis included higher representation from Eastern Mediterranean, Ashkenazi Jewish and Central/South Asian ancestries (Fig. 4d). Of these 141 individuals, deconvolution analysis suggested that 71% have global non-European ancestry contributions of >25% (roughly equivalent to one non-European grandparent; Fig. 4f).

For further down-stream analyses, we completed additional variant-level QC among individuals with European genetic ancestry (based on PCA; n=6,631). We noted HapMap3 variants absent from the VCF were enriched for low minor allele frequency (MAF; Supplementary Fig. 2b), while the vast majority of HapMap3 and cancer PGS variants have higher MAF (Supplementary Fig. 2c). Applying a strict filter for genotype confidence (GP≥0.9 in ≥90% of individuals) retained proportionally more variants with lower MAF (Supplementary Fig. 2d), consistent with reduced imputation certainty at common variants where heterozygous genotypes are more frequent.

Overall, >75% of post-imputation HapMap3 and cancer PGS variants had high-confidence genotype calls (GP≥0.9) in ≥90% of individuals (Fig. 5a). Following variant-level QC (see Methods, Supplementary Fig. 2e-f), we retained >80% of post-imputation HapMap3 and cancer PGS variants. Retention of cancer PGS variants was similar for breast cancer, prostate cancer, and melanoma PGS (70-76% of original variants) and lower for colorectal cancer PGS (62–63% of original variants). Supporting data quality, in the final post-QC dataset for unrelated European-ancestry participants, allele frequencies for retained HapMap3 variants were highly concordant with observed frequencies in the 1000 Genomes European reference population (*r*=0.99, *p*<2.2×10^−16^; Fig. 5b).

**Figure 5.**
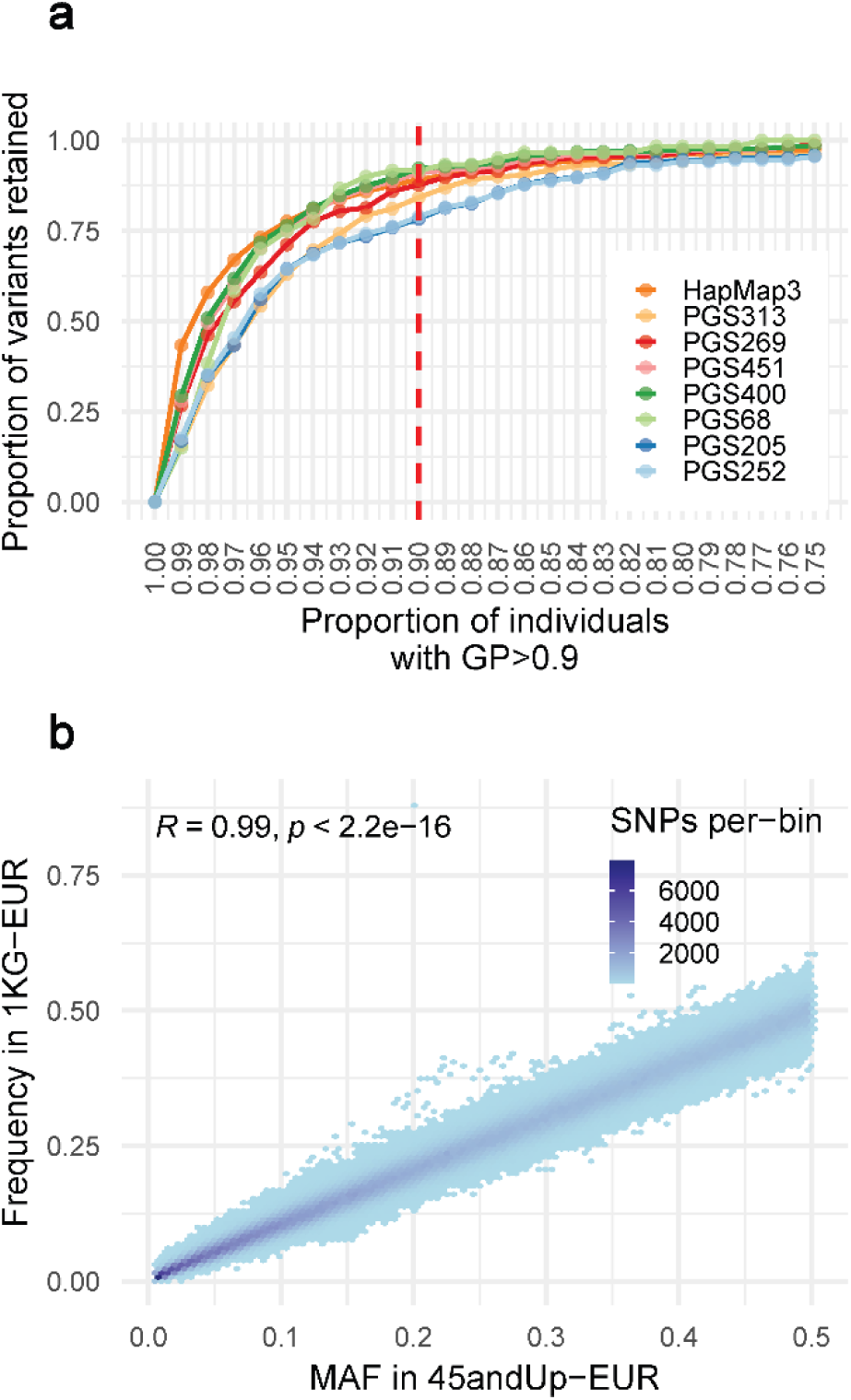
Retained variants by threshold for proportion of individuals with high-confidence (GP ≥0.G) data and final allele frequency concordance with a European reference population, for post-QC data from unrelated European-ancestry participants (n = 6,631). **(a)** Variants retained across different thresholds for minimum proportion of individuals with high-confidence genotype calls (GP≥0.9) among unrelated European-ancestry participants (n = 6,631). Values are shown relative to the total number of post-imputation variants (i.e. variants present in the VCF) for each variant set. The dashed line indicates that >75% of variants have high-confidence genotype calls in ≥90% of individuals. **(b)** Concordance of minor allele frequencies (MAF) among the post-QC unrelated European-ancestry participants and the 1000 Genomes Project European reference population, for post-QC HapMap3 variants. Each hexagonal bin represents the number of variants within a given frequency interval. Allele frequencies are highly concordant between datasets, supporting the reliability of the imputed genotype data.

Ancestry components are grouped according to the Gencove ancestry reference population hierarchy (see Supplementary Information), with abbreviations corresponding to ancestry groups in the Gencove reference panel (see Supplementary Information for details): SWEUROPE–NEEUROPE–NEUROPE, Southwestern, Northeastern, and Northern/Central European ancestry; SCANDINAVIA, Scandinavian ancestry; NNEUROPE, Northern British Isles ancestry; NITALY, Northern Italian ancestry; EMED, Eastern Mediterranean ancestry; ASHKENAZI, Ashkenazi Jewish ancestry; FINLAND, Finnish ancestry; NEASIA–SEASIA–CASIA–EASIA–NCASIA, Northeast Asian, Southeast Asian, Central Asian, East Asian, and North-Central Asian ancestry; TURK–IRAN–CAUCASUS, Anatolian, Caucasus, and Iranian Plateau ancestry; INDPAK–SSASIA, Central and Southern Indian Subcontinent ancestry; SAFRICA–WAFRICA–CAFRICA–EAFRICA–NAFRICA, Southern, Western, Central, Eastern, and Northern African ancestry; NEAREAST, Middle Eastern ancestry; OCEANIA, Oceanian ancestry; AMERICAS, Admixed American ancestry; and BENGALI, Bengali ancestry.

### 2.3 Genotype concordance and reproducibility of lcWGS data

To validate data quality, we assessed genotype concordance using (i) technical duplicate lcWGS samples (duplicate independently sequenced and imputed) and (ii) cross-platform comparisons between lcWGS-derived genotypes and Illumina Global Screening Array (GSA) genotypes generated for a subset of study participants (Fig. 6; Supplementary Fig. 4), considering high-confidence hard-call genotypes (GP≥0.9).

**Figure 6.**
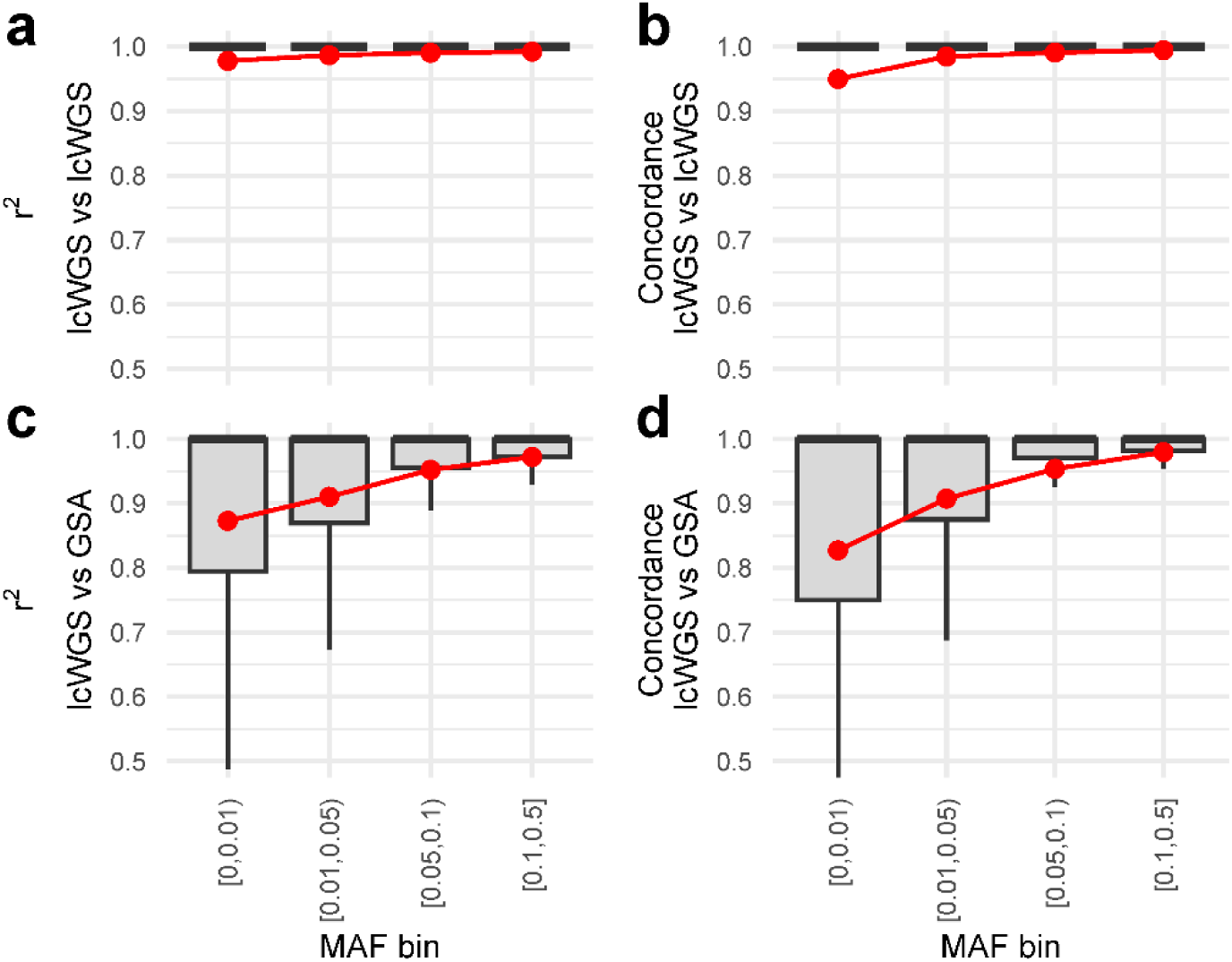
High genotype concordance in comparisons of lcWGS duplicate samples and between lcWGS and dense genotype array data, across variants with different minor allele frequency. **(a–b)** Comparison between duplicate lcWGS sample pairs (n=85 pairs; independently sequenced and imputed) following quality control and restriction to European-ancestry individuals. Genotypes were derived using hard calls based on genotype probability (GP≥0.9), restricted to ∼1.13M HapMap3 and cancer PGS variants. Panel a shows squared genotype correlation (r^2^) and panel b shows minor allele carrier concordance (genotypes 1 or 2) across minor allele frequency (MAF) strata. Raw coverage ratios between duplicate samples exceeded 2.18. **(c–d)** Comparison between lcWGS genotypes and Illumina Global Screening Array (GSA) genotypes for 170 individuals, following quality control and restriction to European-ancestry individuals. lcWGS genotypes were derived using hard calls based on genotype probability (GP≥0.9), including all ∼505K variants shared between imputed lcWGS and directly typed GSA data (not restricted to HapMap3 and cancer PGS variants). Panel c shows squared genotype correlation (r^2^) and panel d shows minor allele carrier concordance across MAF strata. Results were similar when analysing 188 individuals including 18 with non-European ancestry (Supplementary Fig. 4a,b) and when hard-call genotypes in lcWGS data were derived without genotype-probability thresholding (Supplementary Fig. 4c,d).

Across duplicate lcWGS pairs independently sequenced and imputed (n=85 retained post-QC), agreement of genotypes was consistently high for all minor allele frequency (MAF) strata, with near-perfect squared genotype correlation (r^2^) and minor allele carrier concordance (Fig. 6a-b). Despite differences in sequencing depth between duplicates (raw coverage ratio 2.18–2.41), cancer PGS raw values among these duplicate samples also had near-perfect correlation (*r*=0.97–0.99; Supplementary Fig. 5).

In 170 European-ancestry individuals with both lcWGS and Illumina Global Screening Array (GSA) data (n=170), cross-platform concordance was also high across all MAF strata (Fig. 6c-d). As expected, concordance was slightly lower for rare variants (mean r^2^=0.87 for MAF<0.01) and high for common variants (mean r^2^=0.95 for MAF 0.05-0.1, r^2^=0.97 for MAF≥0.1).

Results were similar when analysing all participants with both lcWGS and GSA data (n=188, including 18 non-European ancestry individuals; Supplementary Fig. 4a-b), and when hard-call genotypes in lcWGS data were derived without genotype-probability thresholding, indicating that concordance metrics were highly robust to hard-calling choices (Supplementary Fig. 4c-d).

### 2.4 Evaluation of cancer risk predictions

To showcase the potential for integration of the genetic data with existing 45 and Up Study and linked health data, we completed proof-of-concept evaluations of cancer PGS in this dataset, noting these newly generated data are independent of previous GWAS and PGS analyses. Here, data on cancer diagnoses were derived from population-wide cancer registry records, including age at diagnosis, cancer type, and spread of disease at diagnosis (see Methods). For these analyses, we focused on 6,631 unrelated European-ancestry individuals (Table 2; Supplementary Figs. 6–9). For breast cancer (2,487 female participants, 1,431 breast cancer cases), PGS313^17^ demonstrated a predictive performance of AUC=0.618 (Table 2; Supplementary Fig. 6a), consistent with published validation benchmarks (e.g., AUC 0.60–0.64 in UK and Australian European-ancestry cohorts)^17, 18^. Risk was significantly elevated for individuals in the top 20% of the PGS distribution (OR=1.86; 95%CI: 1.46–2.39) relative to the middle quantile (40–60%), but reduced for those in the lowest 20% PGS (OR=0.54; 95%CI: 0.41–0.72; Supplementary Fig. 7a).

**Table 2.**
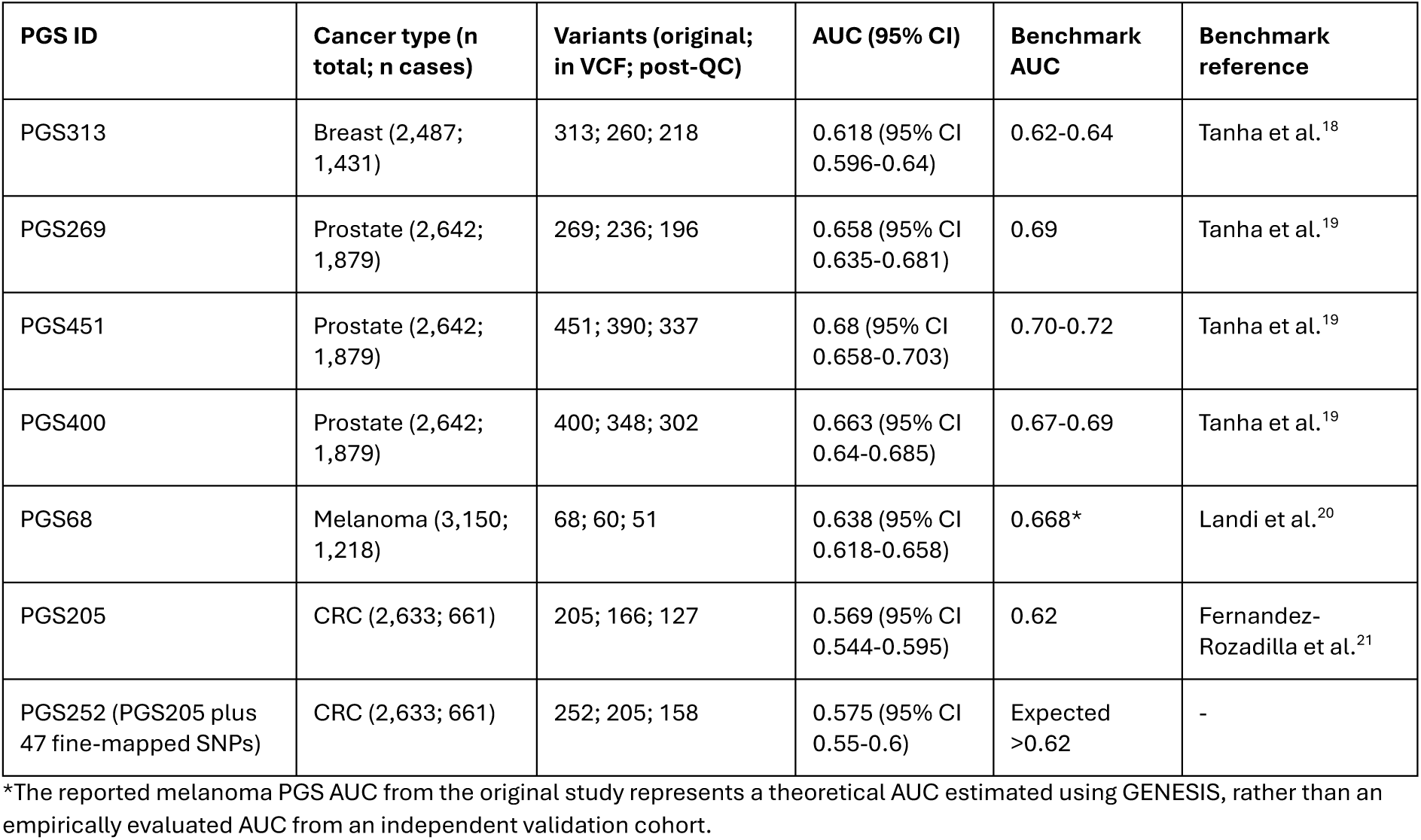
Illustrative independent evaluation of established cancer PGS using newly generated genomic data from unrelated European-ancestry 45 and Up Study participants (n=6,631). For each PGS, the number of variants in the original score, the number present in the imputed VCF, and the number retained after quality control (post-QC) are shown. Discriminative performance is reported as the area under the receiver operating characteristic curve (AUC) with 95% confidence intervals. Benchmark AUC values (where available, in validation studies independent of PGS discovery data) are provided for comparison.

| PGS ID | Cancer type (n total; n cases) | Variants (original; in VCF; post-QC) | AUC (95% CI) | Benchmark AUC | Benchmark reference |
| --- | --- | --- | --- | --- | --- |
| PGS313 | Breast (2,487; 1,431) | 313; 260; 218 | 0.618 (95% CI 0.596-0.64) | 0.62-0.64 | Tanha et al.<sup>18</sup> |
| PGS269 | Prostate (2,642; 1,879) | 269; 236; 196 | 0.658 (95% CI 0.635-0.681) | 0.69 | Tanha et al.<sup>19</sup> |
| PGS451 | Prostate (2,642; 1,879) | 451; 390; 337 | 0.68 (95% CI 0.658-0.703) | 0.70-0.72 | Tanha et al.<sup>19</sup> |
| PGS400 | Prostate (2,642; 1,879) | 400; 348; 302 | 0.663 (95% CI 0.64-0.685) | 0.67-0.69 | Tanha et al.<sup>19</sup> |
| PGS68 | Melanoma (3,150; 1,218) | 68; 60; 51 | 0.638 (95% CI 0.618-0.658) | 0.668* | Landi et al.<sup>20</sup> |
| PGS205 | CRC (2,633; 661) | 205; 166; 127 | 0.569 (95% CI 0.544-0.595) | 0.62 | Fernandez-Rozadilla et al.<sup>21</sup> |
| PGS252 (PGS205 plus 47 fine-mapped SNPs) | CRC (2,633; 661) | 252; 205; 158 | 0.575 (95% CI 0.55-0.6) | Expected >0.62 | - |

\*The reported melanoma PGS AUC from the original study represents a theoretical AUC estimated using GENESIS, rather than an empirically evaluated AUC from an independent validation cohort.

For prostate cancer (2,642 male participants, 1,879 prostate cancer cases), PGS451^22^ yielded an AUC of 0.68 (Table 2; Supplementary Fig. 6b–d), comparable to prior validation estimates (AUC 0.667–0.718 in other European-ancestry cohorts^19^). Individuals in the top 20% of the PGS distribution had approximately three-fold higher odds of prostate cancer relative to the middle quantile (OR=2.46; 95%CI: 1.88–3.22), whereas those in the bottom 20% had substantially lower risk (OR=0.43; 95%CI: 0.31–0.58; Supplementary Fig. 7b–d).

For melanoma (3,150 participants, 1,218 cases), PGS68^20^ yielded an AUC of 0.638 (Table 2; Supplementary Fig. 6e), comparable to a theoretical AUC estimate of 0.668 (based on statistical genetics modelling rather than an independent validation) ^20^, or a prior validation estimate of 0.69 for absolute risk models based on age and sex adjusted by PGS68^23^. Odds ratios for the top PGS quintile were nearly double relative to the middle quintile (OR=1.98; 95%CI: 1.60–2.46), while risk in the lowest quintile was reduced (OR=0.55; 95%CI: 0.42–0.70; Supplementary Fig. 7e).

For colorectal cancer (2,633 participants, 661 cases), PGS205^21^ showed lower predictive performance, with an AUC of 0.569 (Table 2; Supplementary Fig. 6f–g) and little separation between PGS distributions for cases and controls. This AUC was lower than a previously reported estimate of 0.62 (95%CI: 0.60–0.63)^21^. Risk differences were modest: OR=1.36 (95%CI: 1.04–1.76) for the top and OR=0.67 (95%CI: 0.50–0.91) for the lowest quintile, each relative to the middle quintile (Supplementary Fig. 7f-g). PGS252 (PGS205 plus 47 fine-mapped SNPs from Chen et al.^24^) showed only a small improvement (AUC=0.575).

As a sensitivity analysis, we compared PGS performance using all score variants present in the imputed VCF with performance using the post-QC variant set (Supplementary Table 3). Differences in AUC were small across all PGSs. In particular, the two best-performing prostate cancer PGSs (PGS451 and PGS400) showed negligible differences after quality control, supporting the use of the post-QC variant set for subsequent analyses. Although nominally higher AUCs were observed for several other PGSs before QC, these differences were small and not statistically significant.

To illustrate an example of the value of linked health data, we assessed associations between PGS and age at cancer diagnosis among cases (Supplementary Fig. 8). For prostate cancer, there was a highly significant association between younger age at diagnosis and higher PGS (Spearman *ρ* ∼ –0.15, *p* < 6.6×10^−10^), consistent with previous studies^19, 22^. There were no significant associations between PGS and age at diagnosis for the other three cancers. We also evaluated associations between PGS and spread of cancer at diagnosis (Supplementary Fig. 9); while a few comparisons reached nominal significance, we observed no consistent pattern across cancer types or PGS, suggesting no meaningful evidence for an association between the evaluated PGS and spread of disease at diagnosis in the current data.

## 3. Discussion

This study describes the generation, validation, and initial application of genome-wide genetic data within >7,400 participants of the 45 and Up Study^5^, representing the first large-scale integration of genomic and longitudinal health data within this valuable longitudinal cohort.

Using low-coverage whole-genome sequencing (lcWGS) with imputation, we generated high-quality genomic data for 6,827 participants, including 6,631 unrelated European-ancestry individuals. These data establish a scalable genomic data generation framework for one of the largest cohorts in the Southern Hemisphere and provide an important foundation for future genetic, epidemiological, and translational research in Australia and internationally.

Multiple complementary evaluations support the high quality of post-QC lcWGS-derived genetic data. Notably, genotype concordance for independently sequenced duplicate samples was near perfect across the allele frequency spectrum, with minor allele carrier concordance remaining high even for variants with MAF<0.01. Cross-platform comparisons with dense genotype array data also demonstrated high genotype concordance, particularly for common variants. These findings are consistent with previous studies showing that lcWGS combined with lcWGS-tailored modern imputation methods can achieve accuracy comparable to or exceeding dense genotyping arrays even at low sequencing depths^8, 9, 10, 11, 25, 26^, including at ∼0.5× coverage. Together, our results further support lcWGS as a potential strategy for generating genome-wide data in large population cohorts.

A key strength of this study is the availability of detailed baseline data for all individuals invited to provide DNA samples, regardless of subsequent participation. This enabled direct comparison between participants who consented to provide a DNA sample and those who were invited but did not select to participate, allowing for simultaneous adjustment for a broad range of sociodemographic and health-related characteristics. Such analyses are not feasible in stand-alone genetics studies, where information on non-participants is at best highly limited. The capacity to characterise selection into participation using rich, individual-level data strengthens inference from the generated genetics data and provides important insight into patterns of possible participation biases. It also enables future analyses to weight participant data according to a wide range of characteristics. We found that some features associated with under-representation in genomic studies (notably, non-Australian country of birth and dark skin) had significant negative associations with both consent (among invited participants) and inclusion in final genomic data passing all standard quality checks (among all who consented, and among all who provided a DNA sample, see Supplementary Information), providing insights into compounding effects that contribute to under-representation of diverse populations.

The ancestry composition of the genomic dataset broadly reflected the demographic structure of the underlying cohort, with most participants inferred to be of European ancestry. The inferred genetic ancestry was also broadly consistent with self-reported ancestry information, with specific non-European ancestries reported by only 0.6% of participants with inferred European genetic ancestry, compared with 29% of those with inferred non-European or admixed genetic ancestry. Detailed ancestry deconvolution further showed that the European subset was predominantly composed of Northern/Central European and Scandinavian ancestry components, consistent with historical migration patterns within Australia^27^. Although individuals with non-European or admixed ancestry represented a smaller proportion of the cohort, their inclusion remains important for future efforts aimed at improving the diversity and transferability of genomics insights, as the underrepresentation of non-European populations remains a major limitation across human genetics research^28^.

We further demonstrated the utility of the genomic resource through proof-of-concept analyses of cancer polygenic risk prediction. Across breast, prostate, and melanoma cancers, PGS performance was highly consistent with previously reported benchmarks derived from higher-coverage sequencing or genotyping-array datasets. These findings further support that lcWGS-derived genotypes are suitable for clinically relevant PGS estimation in large population cohorts. As a limitation, colorectal cancer PGS performance was lower than expected compared with the original GWAS study^21^. Several factors may contribute to this observation.

First, colorectal cancer PGSs showed a lower proportion of variants included in imputed genotypes, possibly due to differences between the imputation reference panels used in the original GWAS (primarily HRC, 1000 Genomes, and UK10K) and this study (1000 Genomes, HGDP, and gnomAD)^21^. Second, variant retention following quality control was lower compared with the other cancer PGSs, potentially reflecting a greater enrichment of common variants, where the higher frequency of heterozygous genotypes may increase imputation uncertainty. Finally, the number of cases was substantially lower than for other cancers, contributing to more uncertainty in performance; differences in cohort characteristics, case ascertainment, and disease heterogeneity may also contribute to reduced discrimination. Here, the addition of fine-mapped variants in PGS252 modestly improved performance relative to PGS205, suggesting that future optimisation of colorectal cancer PGSs may improve performance in lcWGS-derived datasets such as the one presented here.

The integration of genomic data with linked longitudinal health information enabled additional exploratory analyses beyond standard discrimination metrics. Higher polygenic risk was associated with younger age at diagnosis for prostate cancer, consistent with previous studies reporting stronger genetic susceptibility among earlier-onset disease^19, 22^. In contrast, there was no association between PGS and disease spread at diagnosis. Although exploratory, these analyses illustrate the unique value of embedding genomic data within a deeply phenotyped longitudinal cohort linked to population-wide administrative health datasets.

Several limitations should be considered. First, although lcWGS performs well for common variation, imputation accuracy for rare variants may remain lower, particularly for variants poorly represented in existing reference panels^8, 29^. Second, most downstream analyses were restricted to European-ancestry individuals, reflecting both the ancestry composition of the cohort and the current limitations of available PGSs. Finally, while the current genomic dataset substantially expands the utility of the 45 and Up Study, additional sequencing efforts specifically targeting underrepresented ancestry groups will be important for improving diversity, representativeness, and the broader applicability of future genomic analyses.

Despite these limitations, this study has several major strengths as described above, especially the combination of genome-wide genetic data with extensive behavioural, sociodemographic, and linked health information, which provides a uniquely rich platform for investigating genetic and non-genetic determinants of disease. Going forward, the scale of the cohort, together with its longitudinal design and continued population-wide linkage infrastructure, creates substantial opportunities for future genome-wide association studies, gene–environment interaction analyses, causal inference studies, and evaluation of risk-based prevention strategies. More broadly, this work provides an example for successful application of lcWGS within a large population cohort and supports the growing use of sequencing-based approaches for scalable genomic epidemiology.

In summary, we demonstrate that lcWGS with imputation can be successfully applied at scale within a large population-based cohort to generate high-quality genomic data suitable for downstream analyses, including polygenic risk prediction. The resulting genomic resource substantially expands the research potential of the 45 and Up Study, providing an important foundation for future genomic and precision health research in Australia, and supporting international research in these areas.

## 4. Materials and Methods

### 4.1 Study Cohort and Sample Selection

This work built on the Sax Institute’s 45 and Up Study, a large population-based cohort of 267,357 adults aged ≥45 years residing in New South Wales (NSW), Australia, recruited between 2005 and 2009 through random sampling from the Services Australia Medicare enrolment database (MEDB) with oversampling of people aged 80+ years and those living in regional and remote areas. Approximately 19% of those invited participated, representing around 11% of the NSW population aged ≥45 years^5^.

Cancers and deaths were identified via probabilistic linkage to the NSW Cancer Registry and the NSW Registry of Births, Deaths and Marriages, respectively, by the Centre for Health Record Linkage (https://www.cherel.org.au/). For the purpose of generating genetic data, we invited 30,541 participants to provide a saliva sample for DNA analysis. Participants who provided consent were mailed a Thermo Scientific SpeciMAX Stabilized Saliva Collection Kit (1 mL) for saliva collection. Genomic DNA was extracted using the Chemagic Saliva DNA Kit (CMG-1081) on the Revvity chemagic™ 360 automated platform with magnetic bead technology, following the manufacturer’s instructions. DNA was eluted in 220 μL Tris-HCl buffer, and samples were processed in batches of 23 with one negative control per batch.

The sample selection strategy ensured inclusion of individuals with a history of one of the four major cancers, aligned with the focus of the Australian Cancer Risk Study, alongside a randomly selected sub-cohort as an enduring resource for future research across health and disease areas. Full participant eligibility details are shown in Supplementary Information. We tested the invitation process using a subset of 2,400 invited participants (Supplementary Information), establishing step-wise invitation approaches separately for individuals with available email address and individuals with only postal address available, aiming for a balance of feasibility and optimising uptake. All participants provided written informed consent for provision of saliva samples and DNA analysis, including combination with existing 45 and Up Study data and data linkage; almost all also provided consent for future use of their genomic and health information for health/medical research with relevant ethics approvals (with detailed consent information held by the Sax Institute, who are the data custodians for the 45 and Up Study). Ethics approval for the 45 and Up Study was provided by the University of NSW Human Research Ethics Committee (HC210602). Specific approval for this study was obtained from the NSW Population and Health Services Research Ethics Committee (references 2019/ETH01746 and HREC/14/CIPHS/54).

### 4.2 Sociodemographic Characteristics

Sociodemographic characteristics of individuals with genomic data generated and the full 45 and Up Study cohort were obtained from self-report in the 45 and Up Study baseline questionnaire (2005-2009)^13^, and cancer-related information was obtained from the NSW Cancer Registry. Characteristics of interest included age at baseline, age at sample collection, sex, highest education level attained, private health insurance status, household income, socioeconomic level of place of residence^30^, accessibility/remoteness of place of residence^31^, country of birth, family history of selected cancer types (breast, colorectal, lung, melanoma, ovary, prostate), history of cancer screening (breast, colorectal, prostate), skin colour, ability to tan, time spent outdoors, self-rated health, history of selected conditions (heart disease/stroke, high blood pressure, depression/anxiety, blood clots), body mass index, alcohol consumption, and smoking status (Supplementary Table 1). Cancer characteristics at diagnosis included year, summary spread of disease, and age at diagnosis.

To assess representativeness, we compared sociodemographic and health characteristics of:

- Individuals who were invited and consented to provide a DNA sample, versus individuals who were invited but did consent to provide a DNA sample;
- Individuals whose genomic data was in the high-quality genomic dataset passing all quality checks (see below), versus individuals who consented, but did not have genomic data in the high-quality genomic dataset passing all quality checks (i.e. did not return a DNA sample, or DNA sample was insufficient for low-coverage WGS, or genomic data were generated but did not pass all quality checks).

We calculated counts and percentages, and tested for association using multivariable logistic regression using stepwise forward selection (p<0.10), adjusting for all characteristics included in the model. This analysis was undertaken in SAS v9.4 (SAS Institute Inc., NC, US), with plots generated in RStudio version 2023.03.0 (RStudio Team).

### 4.3 Sequencing, Quality Control, Imputation, and Ancestry Deconvolution

Genomic data were generated using low-coverage whole-genome sequencing (lcWGS) performed by Gencove, Inc.^8^ Sequencing libraries were prepared using a modified protocol based on the Illumina Nextera DNA Flex Library Preparation kit. Final libraries were quantified using fluorometric measurement, and library pools were assessed for fragment quality using the Agilent TapeStation system. Standard Illumina adapter sequences were used for library construction. Sequencing was carried out on an Illumina NovaSeq 6000 platform.

Gencove’s proprietary lcWGS pipeline includes a comprehensive suite of automated initial quality checks (QC) designed to ensure the integrity and analytical suitability of sequence data. Samples that failed any initial QC metric were not included in downstream analysis. Raw sequencing coverage was estimated from the total number of bases sequenced per sample, assuming that 1× coverage corresponds to approximately 3.1×10⁹ bases (haploid genome size). In custom data generation for this study, samples with estimated raw coverage <0.4× (equivalent to <1.32×10⁹ bases sequenced) were excluded in initial QC. In addition, based on standard lcWGS Gencove pipeline criteria, valid samples were required to have up to 19.8×10⁹ bases sequenced (approximately up to 6.4× coverage), avoiding deeper coverage beyond the intended lcWGS design.

Gencove also assessed the number of the reference-panel (described below) variant sites covered by at least one read, as imputation performance depends critically on even genomic coverage (Fig. 2a). For our study, samples were required to have at least 5 million reference-panel SNPs covered by at least one read; values below this threshold indicated insufficiently broad coverage for accurate imputation.

As part of the primary pipeline, Gencove inferred sex chromosome karyotype by calculating the ratio of deduplicated reads mapping to chromosomes X and Y. Samples were classified as XX (female), XY (male), or “Other” when read ratios fell outside empirically validated thresholds. Samples with zero deduplicated mapped reads were labelled “Unknown” due to insufficient information to infer karyotype. A contamination check was performed by examining mitochondrial reads for evidence of multiple mitochondrial haplotypes. Reads deviating from each individual’s consensus mitochondrial profile were used to estimate the proportion of contaminating material. No samples exceeded contamination thresholds.

All samples passing sequencing and QC filters were then processed using Gencove’s genotype imputation workflow, which is based on GLIMPSE2^29^, a method specifically optimised for low-coverage sequencing. The final data included n=7,408 unique individuals, plus a duplicate for n=100 (comprising 91 pairs independently sequenced, genotyped, and imputed, and 9 pairs imputed independently using the same underlying sequencing data). Imputation was performed directly from sequencing reads, rather than from intermediate genotype calls, allowing the algorithm to model sequencing uncertainty and leverage the full information content of lcWGS.

In particular, Gencove’s pipeline uses an adapted implementation of the Li and Stephens (2003)^32^ haplotype-based model, as extended by Rubinacci et al.^29^ for low-coverage data, which delivers fast and accurate imputation over tens of millions of variants. The haplotype reference panel consisted of a merged dataset including the 1000 Genomes Project^14^, Human Genome

Diversity Panel (HGDP)^15^, and gnomAD genomes^16^, aligned to GRCh38. This diverse reference panel provides broad ancestry representation and improves imputation performance across global populations. For each participant and variant, the pipeline outputs genotype probabilities, hard genotype calls, and dosage values in standard variant call format (VCF), enabling flexible downstream analyses including GWAS, PGS, and ancestry inference. Across all samples, the final imputed dataset contained 79,275,947 variants.

To summarise genome-wide imputation performance at the sample level, we calculated phred-scaled missingness, defined as −10 log_10_(*m*), where *m* is the proportion of variants with low-confidence genotype calls (genotype probability GP<0.9). This metric provides an aggregate measure of imputation certainty across the variant set of interest, with higher values indicating lower missingness and thus higher imputation quality. This metric has been used in analogous lcWGS studies and provides a convenient sample-level summary of imputation precision^33^. Following initial QC by Gencove, all samples showed missingness of *m*<0.27. For downstream analyses, we applied a more stringent threshold corresponding to 5% low-confidence variants (*m* ≤0.05; phred ≈ 13).

Finally, Gencove generated global ancestry estimates using an adaptation of the admixture model described by Pritchard, Stephens and Donnelly^34^, modified for sequence-based inputs. Using a supervised framework and a diverse multi-population reference panel, the algorithm estimates each participant’s genome-wide ancestry proportions.

### 4.4 Additional Quality Control Procedures

As an additional quality assessment beyond the QC conducted by Gencove, we implemented a series of sample-level and variant-level quality control procedures to ensure robust downstream analyses. These procedures were performed using PLINK v2.0.0-a6.2^35^ (www.cog-genomics.org/plink/2.0/) and were based on a harmonised variant set comprising GRCh38-lifted HapMap3^36^ SNPs (1,456,478 variants) and SNPs included in several published cancer polygenic scores (PGSs), resulting in a combined variant list of 1,456,478 SNPs, of which 1,334,671 variants (92%) were present in our imputed VCF dataset. The included cancer PGSs were: breast cancer PGS313^17^ (2017 GWAS^37^); prostate cancer PGS269 (2021 GWAS^38^), PGS451 (2023 GWAS^22^) and PGS400 (a version of PGS451 excluding PSA-associated variants); melanoma PGS68 (2020 GWAS^20^); colorectal cancer PGS205 (2023 GWAS^21^) and PGS252 (PGS205 plus 47 fine-mapped SNPs from Chen et al.^24^).

#### 4.4.1 Initial QC, sample verification and correction

Prior to the formal QC steps described below, we first performed a preliminary round of sample-level quality control on the imputed lcWGS dataset (with criteria as described in steps 1-2 below), identifying 6,741 samples that passed and 667 that failed at least one criterion in this preliminary QC. To further assess data integrity, we genotyped all of the 667 latter samples and 120 samples that passed preliminary QC (100 randomly selected with inferred European genetic ancestry, 20 randomly selected with non-European or admixed genetic ancestry), using the Illumina Global Screening Array (GSA; Infinium Global Screening Array-24 v3.0 with multi-disease content add-on), with further details provided below. The array assays approximately 730K variants, including genome-wide backbone content and additional multi-disease markers. Genotype array data were successfully generated (<5% sample-level missingness) for 109 of 120 individuals from the preliminary-QC-passed group and 386 of 667 samples from the preliminary-QC-failed group.

Comparison of lcWGS and GSA genotypes identified label swaps affecting two lcWGS plates (i.e. results from plate *i* were incorrectly labelled as samples on plate *j* and vice versa). Sample identifiers for the affected plates were corrected based on lcWGS vs array genotype matching, and then the formal QC process as described below was completed.

#### 4.4.2 Step 1: Preliminary SNP filtering for sample QC

We first applied stringent filtering to define a high-confidence variant subset for sample-level QC. Variants were required to have genotype probability (GP) ≥0.9, be autosomal, have per-variant missingness ≤0.05 (at GP ≥0.9), minor allele frequency (MAF) ≥0.05, and Hardy–Weinberg equilibrium (HWE) *p* ≥1×10⁻⁶). This filtered SNP set was used exclusively for subsequent sample-level QC.

#### 4.4.3 Step 2: Multi-step sample-level QC

Using the filtered variant set from Step 1, we conducted a multi-step sample-level QC workflow on data for 7,408 individuals plus technical duplicates for 100 individuals. We excluded samples with inconsistencies between reported sex in existing 45 and Up Study epidemiological data and genetically inferred sex; samples with >5% missing genotypes; and individuals with heterozygosity rates outside mean ± 3 standard deviations (SD). For technical duplicate pairs, the sample with the higher call rate was retained.

Pairwise genetic relatedness between all samples was assessed using PLINK v2.0.0-a6.2^35^ with the “--make-king” function, which implements the KING-robust estimator^39^. This method provides kinship coefficients that are robust to population structure and also reports the proportion of opposite homozygotes (IBS0), which aids in distinguishing levels of relatedness. Relatedness filtering was performed using kinship estimates^39^, retaining one individual from each first- or second-degree relative pair and favouring the sample with lower missingness.

Conventional interpretation thresholds were applied: ∼0.354 for monozygotic twins or technical duplicates, ∼0.177 for first-degree relatives (e.g., parent–child, full siblings), ∼0.088 for second-degree relatives (e.g., half-siblings, grandparent–grandchild, avuncular), and ∼0.044 for third-degree relationships.

For ancestry filtering, we projected all individuals onto the first four principal components (PCs) derived from the 1000 Genomes reference data^14^. We retained individuals as European-ancestry if their PC1–PC4 values fell within 3 standard deviations of the mean from 1000 Genomes European data. Heterozygosity, relatedness, and ancestry analyses were performed using the same linkage disequilibrium (LD)-pruned SNP set (500 kb window, r^2^ < 0.1) derived from the Step 1 variant set.

For cross-referencing between inferred genetic ancestry, we also obtained self-reported ancestry from the baseline questionnaire question, “What is your ancestry? (please cross up to 2 boxes)”, for which participants could select zero, one or two responses from Australian, English, Irish, Chinese, Italian, Greek, Scottish, German, Lebanese, Dutch, Maltese, Polish, Filipino, Indian, Croatian, Vietnamese, or ‘Other’. To compare self-reported ancestry with genetically inferred ancestry, we defined two indicators: (1) reporting at least one of Chinese, Filipino, Indian, Lebanese, Maltese or Vietnamese ancestry (the only specific non-Australian and non-European ancestries listed in the questionnaire); and (2) reporting ‘Other’ ancestry (noting this could include less common European ancestries not listed in questionnaire options). The selection of ‘Australian’ ancestry was not deemed informative, as this option was selected by >50% of all 45 and Up Study participants and could reflect any individuals born in Australia who identify as such.

#### 4.4.4 Step 3: Main variant-level QC (restricted to European-ancestry subset)

Sample-level QC retained 6,631 unrelated participants with inferred European genetic ancestry. To support robust downstream polygenic score analyses, we further retained variants that had high-confidence genotype data (GP≥0.9) in ≥90% of participants, which included >75% of variants with available post-imputation data. Additional filters included MAF ≥0.005 and HWE *p* ≥1×10⁻⁶. These thresholds balance stringency with retention of informative variants for PGS evaluation.

Allele frequency concordance was assessed by calculating the Pearson correlation between MAFs of post-QC HapMap3 variants in the 6,631 European-ancestry 45 and Up Study participants and European samples from the 1000 Genomes Project^14^.

### 4.5 Genotype concordance and validation

Genotype concordance and reproducibility were evaluated using two complementary approaches. First, we considered duplicate lcWGS samples (n=85 pairs remaining after QC) that were independently sequenced and imputed. Raw coverage ratios between duplicate samples exceeded 2.18.

Additionally, we used independent genotyping generated via the Illumina Global Screening Array (GSA) with multi-disease drop-in (GSA MD v3) to evaluate cross-platform genotype concordance with lcWGS data. As noted above, genotype arrays were run for 667 individuals with data not passing preliminary QC and 120 individuals with data passing preliminary QC. The arrays were scanned on an Illumina iScan system; the raw fluorescence intensity data was normalized and clustered for each sample using Illumina GenomeStudio v2.0.5 software with the PLINK Plug-in v2.1.4, with data for samples passing call rate >95% clustered separately to call genotypes for these samples.

We thus considered GSA genotype data for 495 individuals with lcWGS data available. An initial comparison identified a label swap for two lcWGS sequencing plates (i.e. results from plate *i* were incorrectly labelled as samples on plate *j* and vice versa). Following correction of relevant sample labels and completion of the full lcWGS QC process as described above, GSA data were available for 188 individuals included in the final high-quality lcWGS dataset, including 170 individuals of inferred European ancestry; these individuals were used for cross-platform genotype concordance analyses.

For individuals with high-quality lcWGS data and GSA genotype data, agreement was quantified using squared Pearson correlation (r^2^) and minor allele carrier concordance across minor allele frequency bins. These metrics were calculated using ∼1.13M variants for lcWGS–lcWGS comparisons (HapMap3 and cancer PGS variants), and ∼505K variants for lcWGS–GSA comparisons (all variants shared between the lcWGS and GSA datasets, not restricted to HapMap3 and cancer PGS).

### 4.6 Polygenic Risk Score Calculation

Across the imputed dataset, the number of variants present and retainable in the VCF for each published polygenic score (PGS) is shown in Table 2. Notably, the imputed genotypes included in VCF files only included a subset of variants for each of the PGS: PGS313 (breast cancer) - 260 of 313 variants (83%), PGS269 (prostate cancer) - 236 of 269 variants (88%), PGS451 (prostate cancer) - 390 of 451 variants (86%), PGS400 (prostate cancer; PSA-associated variants excluded) - 348 of 400 variants (87%), PGS68 (melanoma) - 60 of 68 variants (88%), PGS205 (colorectal cancer) - 166 of 205 variants (81%), PGS252 (colorectal cancer; with additional fine-mapped variants): 205 of 252 variants (81%). For PGS calculation, only variants that also passed the main variant-level QC filters described above were retained, resulting in smaller final SNP sets for each PGS (Table 2; e.g., for PGS313, 260 variants were present in the VCF, of which 218 passed all QC filters and were used in scoring; full counts are presented in the Results section).

Raw PGS values for all individuals were computed from genotype dosages using PLINK v2.0.0-a6.2^35^ (the “--score” function). For each score, we used the “SCORE1_AVG” output, which corresponds to the weighted sum of allelic dosages averaged over the number of non-missing allele observations. Raw PGS values were standardized to Z-scores (PGS_Z_) by subtracting the mean and dividing by the standard deviation of a randomly selected sub-cohort.

To assess PGS reproducibility, we evaluated the concordance of raw PGS values for 85 duplicate sample pairs (post-QC) that were independently genotyped and imputed, using Pearson correlation.

Discrimination of each PGS_Z_ for its corresponding cancer outcome was evaluated using the area under the receiver operating characteristic curve (AUC), which reflects the probability that a participant who develops cancer has a higher score than one who does not. An AUC of 0.5 indicates no discriminatory ability (random ranking), while an AUC of 1.0 represents perfect discrimination. AUC estimates, along with 95% confidence intervals (CIs), were calculated using the “pROC” R package with 2,000 stratified bootstrap samples (via the “ci.auc” function). As a sensitivity analysis, AUCs based on all score variants present in the imputed VCF were compared with the primary analysis based on the post-QC variant set.

To evaluate stratification across the PGS distribution, odds ratios (ORs) for cancer were estimated across five PGS_Z_ quantiles, using the 40–60% quantile as the reference category. Logistic regression models were used to obtain ORs and 95% confidence intervals for each quantile.

To showcase the use of linked data, we examined associations between each PGS_Z_ and age at cancer diagnosis among cases using Spearman correlation. Additionally, for individuals with available data on cancer spread (summary stage) at diagnosis, we assessed whether PGS differed by disease spread. Specifically, we compared PGS_Z_ values across cancer stage at diagnosis, categorised as localised (L), regional spread to adjacent organs (R1), regional spread to lymph nodes (R2), and distant metastases (M), using the Wilcoxon rank-sum test. Available case numbers included 1,383 for breast cancer, 1,442 for prostate cancer, 1,163 for melanoma, and 597 for colorectal cancer.

## 5. Data Availability

Study data are available from the data custodians (Sax Institute for the 45 and Up Study and genomic data; NSW Ministry of Health for NSW Cancer Registry and NSW Registry of Births, Death and Marriages) for approved research projects. However, direct provision of the data by the authors is not permitted by the relevant data custodians, as it would compromise participants’ confidentiality and privacy. Data access enquiries can be made to the Sax Institute by email, or see https://www.saxinstitute.org.au/solutions/45-and-up-study/use-the-45-and-up-study for details.

Upon publication, all newly generated genomic data will also be deposited to the European Genome-phenome Archive (EGA).

## 6. Ethical Approval

Ethics approval for the 45 and Up Study was provided by the University of NSW Human Research Ethics Committee (HC210602). Specific approval for this study was obtained from the NSW Population and Health Services Research Ethics Committee (references 2019/ETH01746 and HREC/14/CIPHS/54).

## 7. Author Contributions

Conceptualization: HMT, DG, MM, AEC, NGM and JS; Data curation: HMT and DG; Formal analysis: HMT and DG; Funding acquisition: AEC, NGM, MM and JS; Investigation: HMT, DG, RP, NG and EdG; Methodology: HMT, DG, RP, EdG, GD, KB, AC, AEC, NGM and JS; Supervision: NGM and JS; Validation: HMT, RP, EdG, NGM, JS; Visualization: HMT and DG; Writing – original draft: HMT, DG and JS; Writing – review and editing: All authors.

## 9. Acknowledgements

The authors acknowledge the technical infrastructure and support provided by the Secure Unified Research Environment (SURE), operated by the Sax Institute. We thank the Centre for Health Record Linkage for performing the linkage and the data custodians for providing their data. This research was completed using data collected through the 45 and Up Study (www.saxinstitute.org.au). The 45 and Up Study is managed by the Sax Institute in collaboration with major partner Cancer Council NSW and the NSW Ministry of Health. We sincerely thank the many thousands of participants who continue to contribute to the 45 and Up Study. We are also grateful to Miguel Rentería for his valuable advice on approaches to participant consent; Rebekah Cicero for their assistance in preparing thousands of saliva collection kits for dispatch; Simone Cross for coordinating and managing sample transfers; Hans Luc and Katie Armstrong for their ongoing valuable advice, support, and contributions throughout this project.

