## Supplementary Information for "A Genome-wide Genetic Data Resource for the 45 and Up Study"

### 1. Supplementary Text

#### 1.1. Eligibility criteria

To identify participants who could be invited to consent and provide a DNA sample, we considered all participants alive as of October 2021, and excluded the following:

- Notification of withdrawal, death, or no further contact
- Contact address unavailable / interstate / overseas
- Part of other recent sub-studies
- Previously indicated unwilling to provide saliva sample for research

We issued invitations to 9,986 randomly selected participants who were not excluded based on the above, and all remaining participants with a NSW Cancer Registry record of invasive breast, prostate, melanoma, or colorectal cancer (using International Classification of Diseases 10$^{th}$ edition, topography codes C50, C61, C43, C18-C20, respectively).

#### 1.2. Invitations

In a pilot set of 2,400 invitations (n=1,600 randomly selected, n=200 additional for each cancer type), we tested two models of consent: “fine-grained” and combined “broad” consent (separately or combined for the Australian Cancer Risk Study and future health/medical research), with each invitation including one of the two consent models (50:50 allocation). We found highly similar consent rates (<2% difference) for “fine-grained” and combined “broad” consent, thus proceeded with the combined “broad” consent model for remaining invitations.

Specifically, fine-grained consent was separately for this research project and future approved health and medical research:

PART A. Consent for this research project

I ________________________________________ (please print name) hereby freely consent to take part in the research project *Australian Cancer Risk Study: Genomic risk factors for cancer and cancer risk prediction in the 45 and Up Study*, including providing a small saliva sample (2mL).

PART B. Additional consent for use of genetic information and DNA sample for future approved health and medical research studies

**I also consent** for my genetic data and DNA sample being made available to other authorised researchers in the future for approved health and medical research.

Broad consent was combined for this research project and future approved health and medical research:

Consent for this research project and for use of genetic information and DNA sample for future approved health and medical research studies

I ________________________________________ (please print name) hereby freely consent to take part in the research project *Australian Cancer Risk Study: Genomic risk factors for cancer and cancer risk prediction in the 45 and Up Study*, including providing a small saliva sample (2mL). I also consent for my genetic data and DNA sample being made available to other authorised researchers in the future for approved health and medical research.

We further used this pilot set to evaluate consent rates at different steps in the invitation process, finding the following consent rates:

- Initial email invitation: 16% (responding within 3 weeks of initial email invitation)
- First email reminder: another 11% (within 5 weeks following a first email reminder, and within total 8 weeks of initial email invitation)
- Second reminder:
  - email: another 11%
  - postal (instead of email): another 9%

All as of interim assessment within 11 weeks of initial invitations, with total consent rate 31% at the interim assessment (11 October 2022), rising to 35% among these participants by the end of the study.

The final invitation process after the pilot phase was as follows.

For 11,776 participants with email address available (including 3,145 randomly selected):

- Initial invitation by email;
- Reminder for invitation by email;
- Reminder for invitation by post;

with an average consent rate of 39%.

For 16,365 participants with no email address available (including 5,241 randomly selected):

- Invitation by post;
- Reminder for invitation by post;

with an average consent rate of 17%.

Detailed consent rates are shown in the following table.

| **Participant group** | **Pilot phase** | **Main invitations, email address available** | **Main invitations, only postal address available** | **Any** |
|---|---|---|---|---|
| Breast cancer history | 45% | 40% | 18% | 29% |
| Prostate cancer history | 40% | 40% | 19% | 29% |
| Melanoma history | 33% | 41% | 18% | 28% |
| Colorectal cancer history | 39% | 37% | 15% | 24% |
| Randomly selected sub-cohort, no history of the four cancers above | 32% | 38% | 16% | 25% |
| All participants | 35% | 39% | 17% | 27% |

Consent rates for different participants groups and by invitation mode. Participants with multiple cancer diagnoses were assigned to the first cancer type diagnosed.

##### 1.3. Gencove ancestry deconvolution reference populations

Ancestry deconvolution was performed using the Gencove reference panel. The reference populations were grouped as follows based on https://resources.gencove.com/hc/en-us/articles/7267880409883-Human-Ancestry-Reference-Populations.

[1] Africa (AFRICA) included Southern Africa (SAFRICA; Ju|'hoansi), Western Africa (WAFRICA; Esan and Yoruba), Central Africa (CAFRICA; Mbuti), Eastern Africa (EAFRICA; Datog, Luhya, Kikuyu and Masai), and Northern Africa (NAFRICA; Algerian, Tunisian, Sahrawi and Mozabite).

[2] Americas (AMERICAS) included Bolivian, Karitiana, Mayan, Mixe, Mixtec, Piapoco, Pima, Surui and Zapotec populations.

[3] Anatolia, Caucasus and Iranian Plateau (TURK-IRAN-CAUCASUS) included Ossetian, Turkish, Abkhasian, Adygei, Armenian, Balkar, Chechen, Druze, Georgian, Iranian, Kumyk and Lezgin populations.

[4] Ashkenazi Jewish (ASHKENAZI) was represented by an Ashkenazi Jewish reference population.

[5] Asia included Northeast Asia (NEASIA; Daur, Hezhen, Mongola, Oroqen, Tu, Ulchi and Xibo), Southeast Asia (SEASIA; Kinh, Dai and Lahu), Central Asia (CASIA; Hazara, Uygur, Uzbek and Kalash), East Asia (EASIA; Han, Miao, Naxi, She, Tujia and Yi), and North-Central Asia (NCASIA; Kalmyk, Tuvinian, Yakut and Altaian).

[6] Bengal (BENGALI) was represented by a Bengali reference population.

[7] Eastern Mediterranean (EMED) included Malta, Sicily, Cyprus, Greece, Albania and Bulgaria.

[8] Europe included Southwestern Europe (SWEUROPE; Southern French, Spanish, Basque and Sardinian), Northeast Europe (NEEUROPE; Belarusian, Estonian, Lithuanian, Mordovian, Finnish, Russian and Ukrainian), and Northern and Central Europe (NEUROPE; British, Irish and German).

[9] Finland (FINLAND) was represented by a Finnish reference population.

[10] Indian Subcontinent included Central Indian Subcontinent populations (INDPAK; Gujarati, Sindhi, Pathan, Burusho, Balochi, Makrani and Brahui) and Southern Indian Subcontinent populations (SSASIA; Telugu and Tamil).

[11] Middle East (NEAREAST) included Bedouin, Egyptian, Jordanian, Palestinian, Saudi, Syrian and Yemeni populations.

[12] Northern British Isles (NNEUROPE) included British, Orcadian and Scottish populations.

[13] Northern Italy (NITALY) included Tuscan and Northern Italian populations.

[14] Oceania (OCEANIA) included Native Australian and Papuan populations.

[15] Scandinavia (SCANDINAVIA) included Icelandic, Norwegian and Swedish populations.

#### 1.4. Participant characteristics associated with genomic data being included in the high-quality dataset passing all quality checks

We investigated characteristics associated with genomic data being included in the high-quality dataset passing all quality checks (n=6,827 individuals, prior to any exclusions based on relatedness or inferred genetic ancestry), among all people who consented to provide a DNA sample. The results are shown in Supplementary Figure 10.

There were significantly ($p$<0.001) lower odds of inclusion in the final high-quality genomic dataset for males (aOR=0.75 (95%CI: 0.66-0.85) versus females), those with dark olive/brown/black skin (aOR=0.46 (95%CI: 0.32-0.67) versus very fair), and those born outside Australia (aOR=0.77 (95%CI: 0.67-0.88)). Inclusion in the final high-quality genomic dataset was also less likely (p<0.01) for those who smoked cigarettes at baseline (aOR=0.64 (95%CI:0.49-0.84)). By contrast, inclusion in the final high-quality genomic dataset was positively associated (p<0.01) with reporting previous cancer screening at baseline (aOR=1.36 (95%CI:1.10-1.68)), drinking >15 alcoholic drinks per week at baseline (aOR=1.31 (95%CI:1.08-1.59) versus 1-5 drinks per week) and household income of $50,000-$70,000 at baseline (aOR=1.35 (95%CI:1.09-1.67) versus <$30,000)).

#### 2. Supplementary Figures

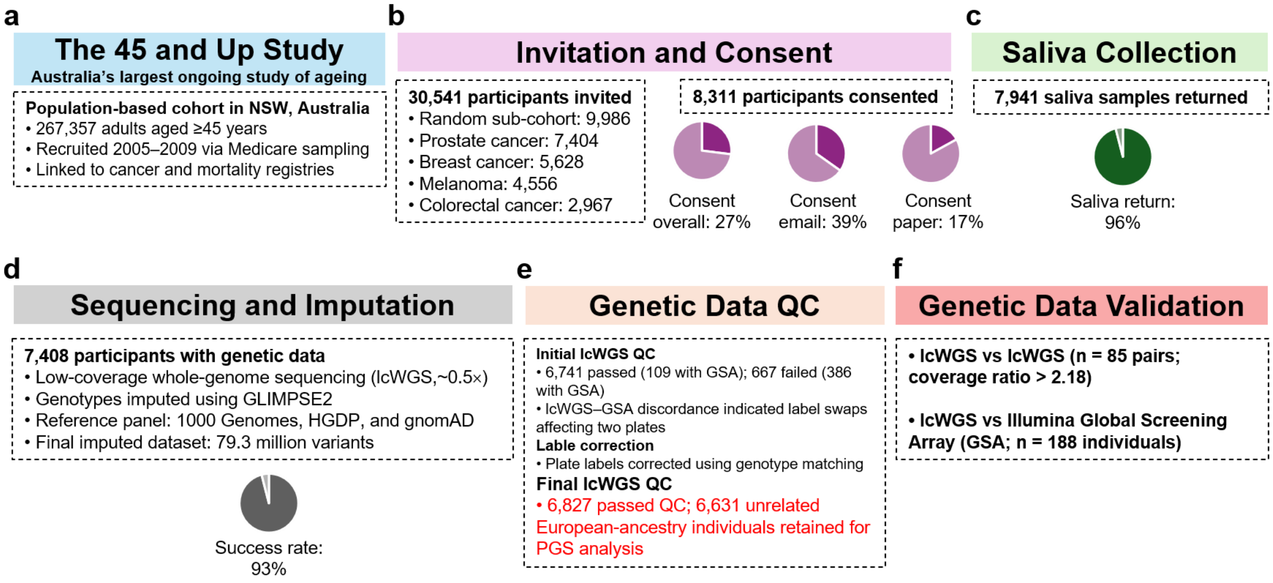

**Supplementary Figure 1. Overview of participant recruitment, genomic data generation, quality control, and validation in the 45 and Up Study sub-cohort. (a)** The 45 and Up Study is Australia's largest ongoing study of ageing, comprising 267,357 participants aged ≥45 years recruited between 2005 and 2009. **(b)** Invitation and consent process for the genomic sub-study. A total of 30,541 participants were invited, including a randomly selected sub-cohort (n = 9,986) and individuals diagnosed with prostate, breast, melanoma, or colorectal cancer who were alive as of October 2021. Overall, 8,311 participants consented to genetic testing and data linkage (27% overall consent rate; 39% among email invitations and 17% among paper invitations). **(c)** Saliva sample collection. Of those who consented, 7,941 saliva samples were returned (96% response rate). **(d)** Sequencing and imputation. Genomic data were generated for 7,408 participants using low-coverage whole-genome sequencing (lcWGS; >0.4X coverage). Genotypes were imputed using GLIMPSE2 with a reference panel combining the 1000 Genomes Project, HGDP, and gnomAD (GRCh38), yielding an imputed dataset of approximately 79.3 million variants. **(e)** Genomic data quality control (QC) workflow. Standard sample-level QC filters were applied, including checks for sex concordance, genotype missingness, and heterozygosity. After initial QC, 6,741 samples passed and 667 failed. Illumina Global Screening Array (GSA) data were successfully generated for a subset of samples (109 QC-passed and 386 QC-failed). Discordance between lcWGS and GSA data identified sample mismatches consistent with label swaps affecting two lcWGS plates. Sample identifiers were corrected based on genotype matching, and QC was repeated, resulting in 6,827 samples passing QC. Of these, 6,772 were unrelated individuals, including 6,631 of European genetic ancestry retained for polygenic score analyses. **(f)** Genomic data validation. Genotype concordance and reproducibility were evaluated using duplicate lcWGS sample pairs (n = 85) and cross-platform comparison with Illumina Global Screening Array (GSA) genotypes (n = 188 individuals).

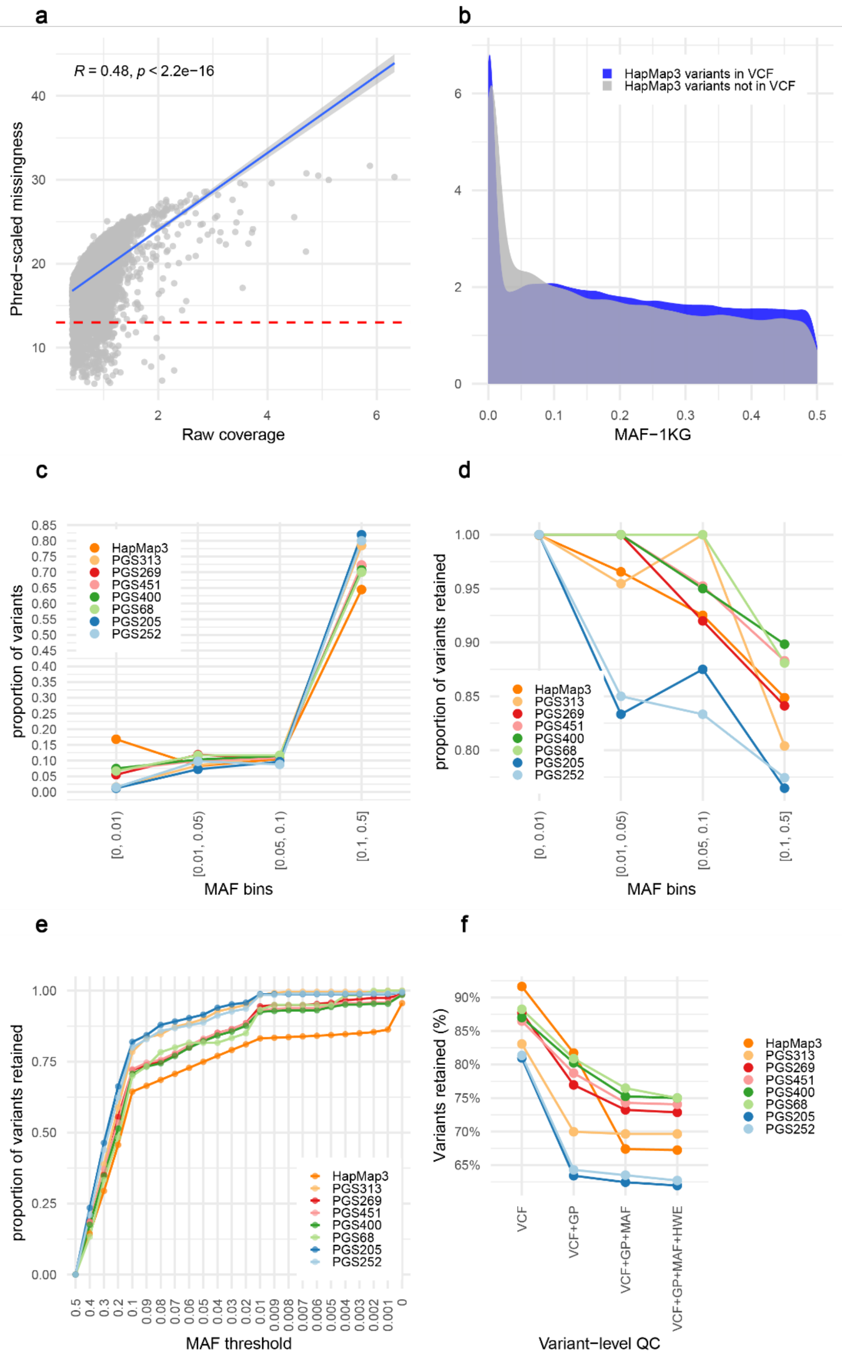

**Supplementary Figure 2. Coverage, missingness, and variant-level quality control of lcWGS data. (a)** Association between raw sequencing coverage and phred-scaled genotype missingness (defined as the proportion of variants with genotype probability GP<0.9), based on all lcWGS data (7,408 unique individuals plus 100 duplicate samples). The dashed line indicates the threshold corresponding to 5% low-confidence genotype calls (phred ≈ 13), equivalent to approximately 0.43X raw coverage. **(b)** Distribution of minor allele frequencies (MAF) for HapMap3 variants present or absent in the post-imputation VCF, based on 1000 Genomes data. Variants that are not present in the VCF are enriched at lower allele frequencies. **(c)** Distribution of HapMap3 and polygenic score (PGS) variants across MAF bins for variants present in the post-imputation VCF, among 6,631 unrelated individuals of European genetic ancestry. **(d)** Proportion of variants contained in the post-imputation VCF that were retained across MAF bins after applying the genotype confidence filter (GP≥0.9 in ≥90% of individuals), based on 6,631 unrelated European-ancestry individuals. **(e)** Proportion of variants retained across MAF thresholds among 6,631 unrelated European-ancestry individuals. **(f)** Variant-level quality control for data from 6,631 unrelated European-ancestry individuals. Sequential filtering steps were applied based on genotype probability (GP), per-variant missingness

(≤0.1, i.e. GP≥0.9 for ≥90% of individuals), minor allele frequency (MAF ≥0.005), and Hardy–Weinberg equilibrium (HWE $p \geq 1\times10^{-6}$). The proportion of variants retained after each step is shown for HapMap3 variants and variants included in published PGS for breast (PGS313), prostate (PGS269, PGS451 and PGS400), melanoma (PGS68), and colorectal (PGS205 and PGS252) cancers.

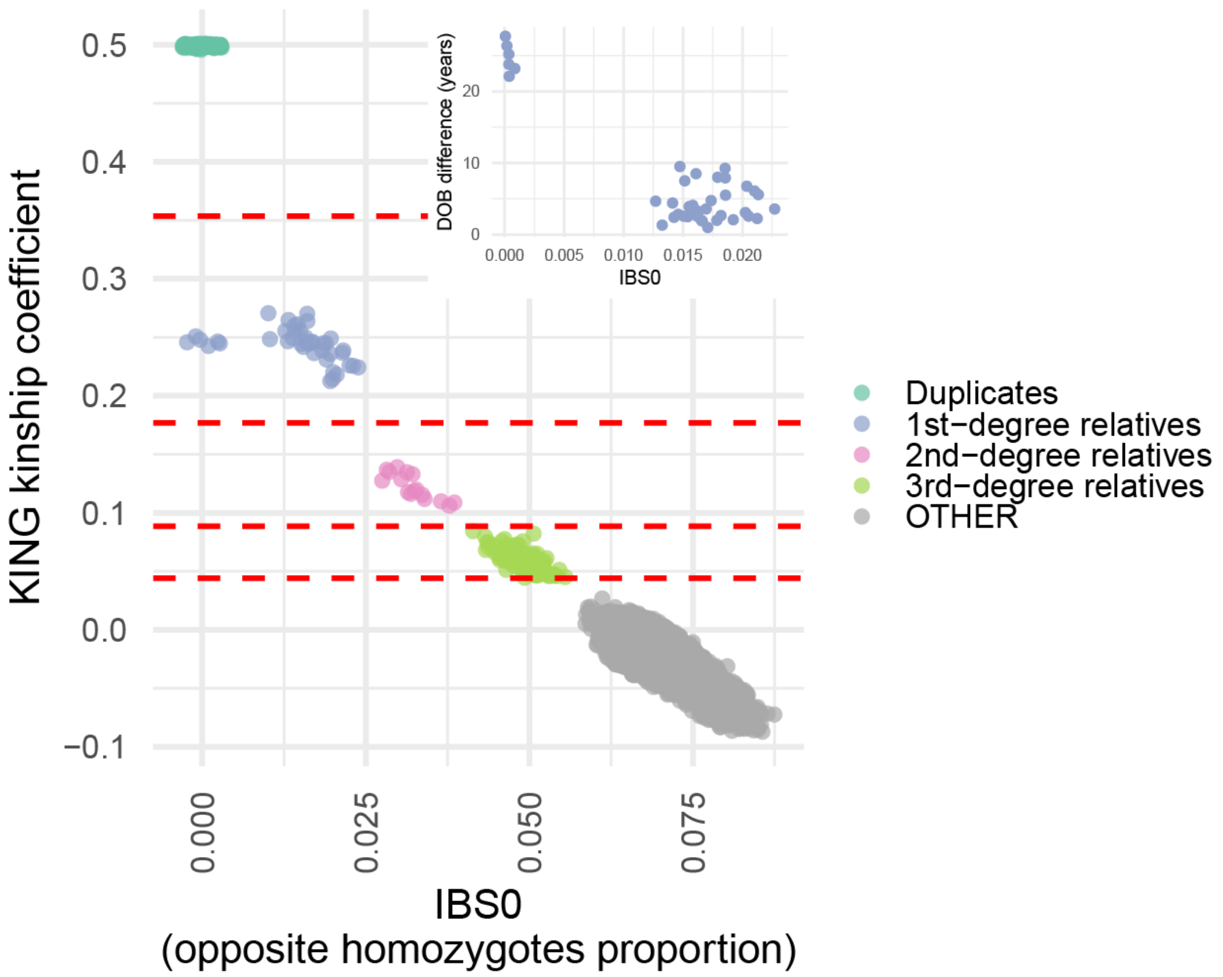

**Supplementary Figure 3. Pairwise genetic relatedness among individuals with genomic data.** Pairwise relatedness was estimated using the KING-robust estimator based on genotype data from 6,921 samples that passed sample-level quality control for sex concordance, sample missingness ≤5%, and heterozygosity within 3 s.d. of the mean. Kinship coefficients are plotted against the proportion of opposite homozygotes (IBS0), enabling discrimination between degrees of relatedness. Duplicate samples and monozygotic (MZ) twins cluster at kinship coefficients of ~0.5 with IBS0 ≈ 0, including 85 duplicate pairs genotyped and imputed independently; 9 duplicate pairs imputed independently; and <5 MZ twin pairs (number suppressed to preserve confidentiality). First-degree relatives cluster around kinship ≈ 0.25, with IBS0 distinguishing parent–child pairs (IBS0≈0) from full siblings (IBS0>0), supported by differences in date of birth (DOB). Second- and third-degree relatives form additional clusters at lower kinship values. For downstream analyses, one sample from each duplicate pair and one individual from each first- or second-degree relative pair was removed, prioritising retention of samples with lower genotype missingness (based on GP≥0.9).

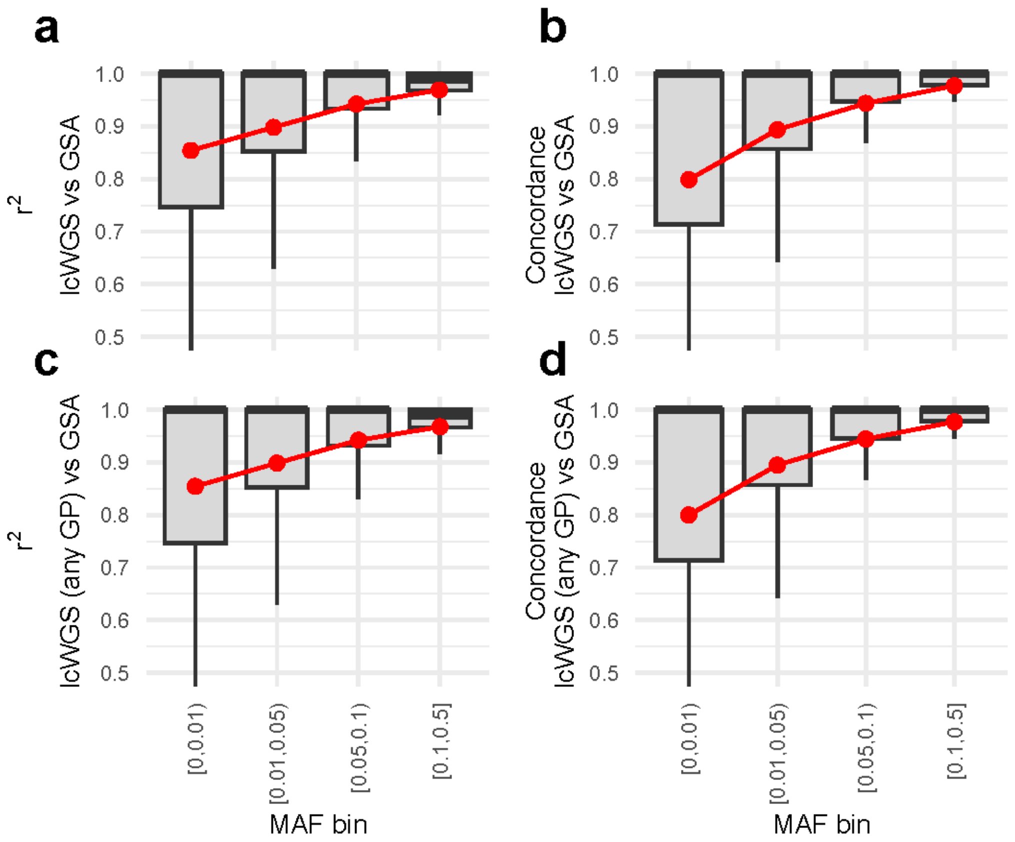

**Supplementary Figure 4. High genotype concordance in comparisons between lcWGS and dense genotype array data – additional analyses including 188 participants. (a–b)** Comparison between lcWGS genotypes and Illumina Global Screening Array (GSA v3 with multi-disease add-on) genotypes for 188 individuals following quality control (170 European-ancestry and 18 non-European or admixed ancestry), with lcWGS genotypes derived using hard calls based on genotype probability (GP≥0.9). Panel a shows squared genotype correlation ($r^2$) and panel b shows minor allele carrier concordance across minor allele frequency (MAF) bins. **(c–d)** Comparison between lcWGS genotypes and Illumina Global Screening Array (GSA v3 with multi-disease add-on) genotypes for 188 individuals following quality control (170 European-ancestry and 18 non-European ancestry), with lcWGS genotypes derived using hard calls without applying a genotype probability threshold (i.e. using all genotype calls). Panel c shows squared genotype correlation ($r^2$) and panel d shows minor allele carrier concordance across MAF bins. All panels show results based ~505K variants shared between imputed lcWGS and directly typed GSA data (not restricted to HapMap3 and cancer PGS variants).

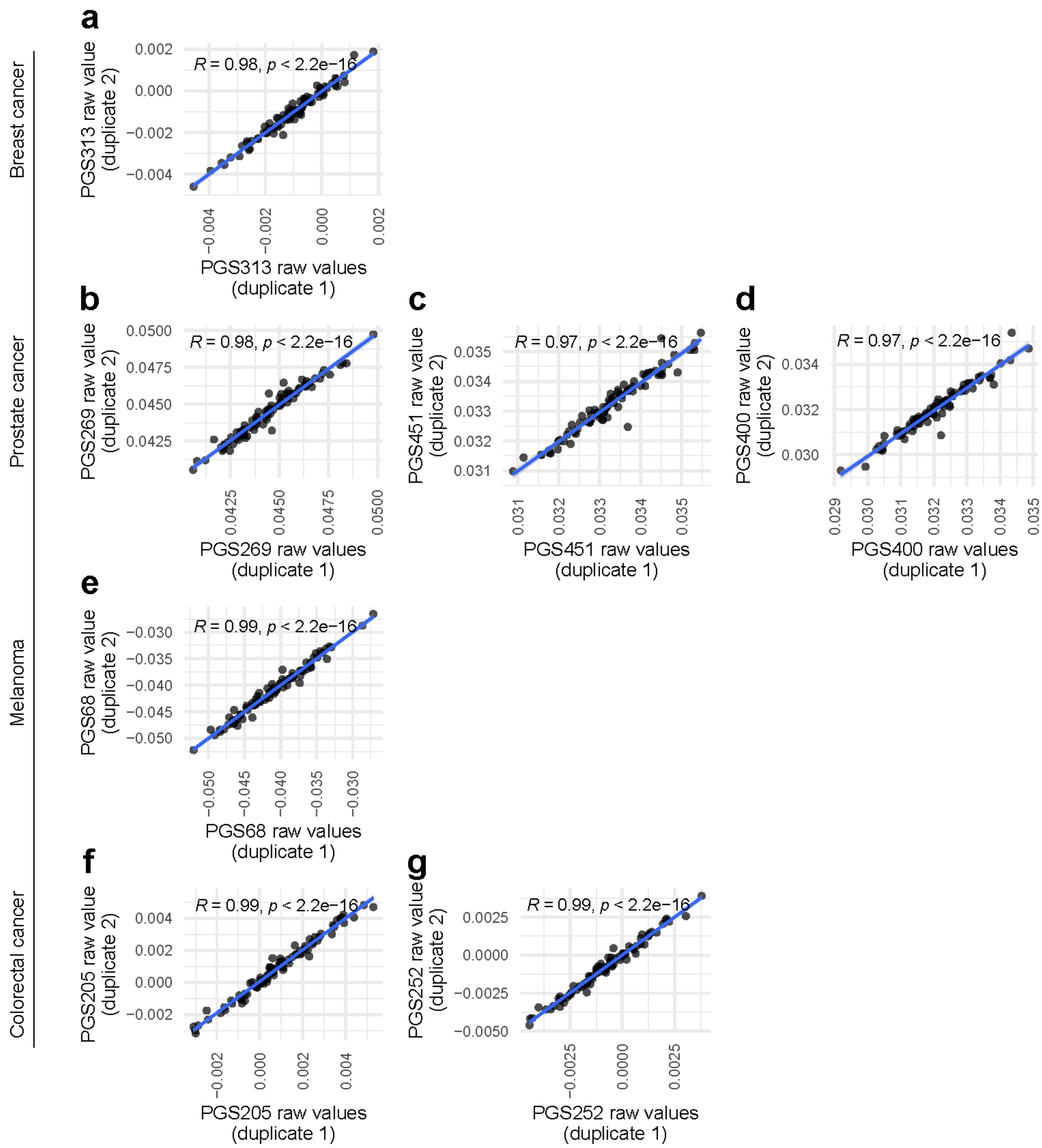

**Supplementary Figure 5. Extremely high correlation between cancer polygenic scores calculated in duplicate lcWGS samples.** Polygenic scores (PGSs) were calculated independently for 85 duplicate sample pairs (independently sequenced and imputed) following quality control and restriction to individuals of European genetic ancestry, with each pair sequenced and imputed separately. **(a–g)**, Comparison of raw PGS values between duplicate samples for different cancer-specific PGSs, including breast cancer (**a**, PGS313), prostate cancer (**b–d**, PGS269, PGS451 and PGS400), melanoma (**e**, PGS68), and colorectal cancer (**f,g**, PGS205 and PGS252). Points represent individual duplicate pairs, and the blue line indicates the linear regression fit. Pearson correlation coefficients (R) and associated P values are shown in each panel. Raw sequencing coverage ratios between duplicate samples (duplicate 2 vs duplicate 1) ranged from 2.18 to 2.41.

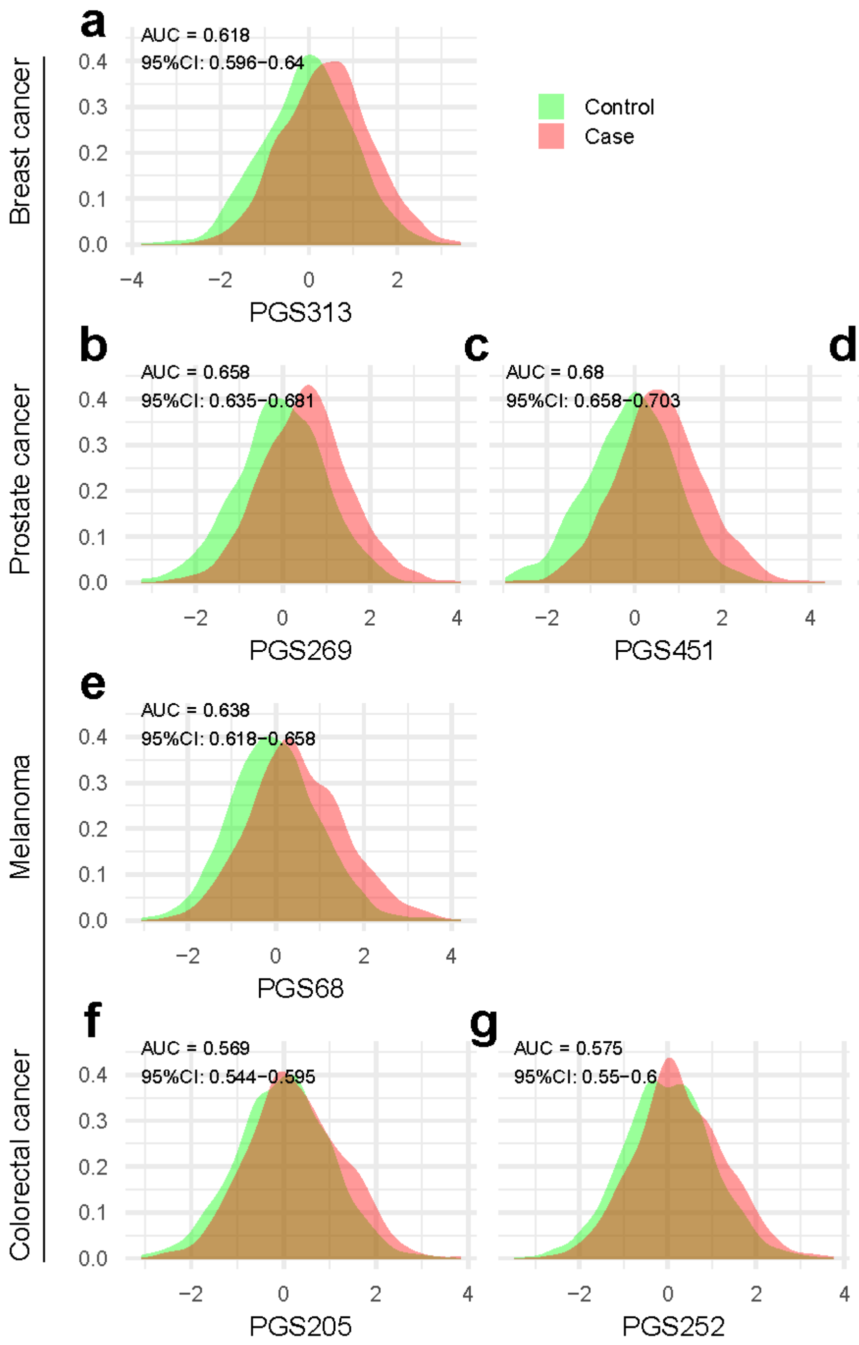

**Supplementary Figure 6. Cancer polygenic score distributions in individuals with diagnosis of that cancer ("cases") and individuals in the randomly selected sub-cohort without a diagnosis of that cancer ("controls"). (a–g)** Distributions of standardised polygenic score (PGS) values (z-scores) in cases and controls for breast cancer (a, PGS313), prostate cancer (**b–d**, PGS269, PGS451 and PGS400), melanoma (**e**, PGS68), and colorectal cancer (**f,g**, PGS205 and PGS252). Area under the receiver operating characteristic curve (AUC) values with 95% confidence intervals are shown in each panel. All analyses were restricted to unrelated European-ancestry individuals; for breast cancer and prostate cancer, analyses were further restricted to female and male participants only. Numbers of included participants are shown in Table 2.

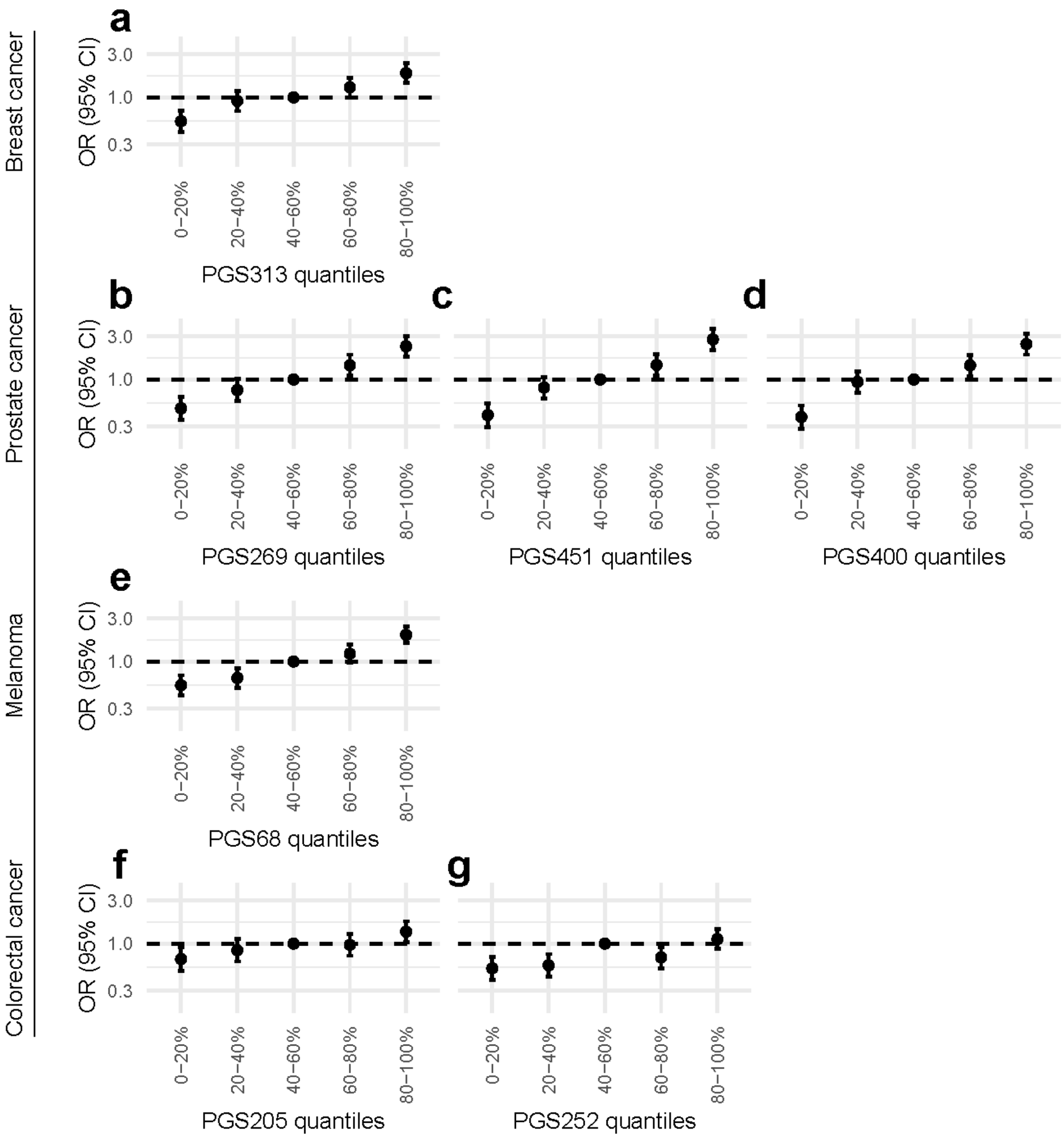

**Supplementary Figure 7. Illustrative analysis of cancer polygenic risk scores (PGS): odds ratios for cancer diagnosis by PGS quantile. (a–g)** Odds ratios (ORs) and 95% confidence intervals for cancer risk across polygenic score (PGS) quantiles, relative to the middle quantile (40–60%). Results are shown for breast cancer (a, PGS313), prostate cancer (**b–d**, PGS269, PGS451 and PGS400), melanoma (**e**, PGS68), and colorectal cancer (**f,g**, PGS205 and PGS252). All analyses were restricted to unrelated European-ancestry individuals; for breast cancer and prostate cancer, analyses were further restricted to female and male participants only. For each cancer, analyses included participants with a diagnosis of that cancer ("cases") and participants in the randomly selected sub-cohort without a diagnosis of that cancer ("controls"). Numbers of included participants are shown in Table 2.

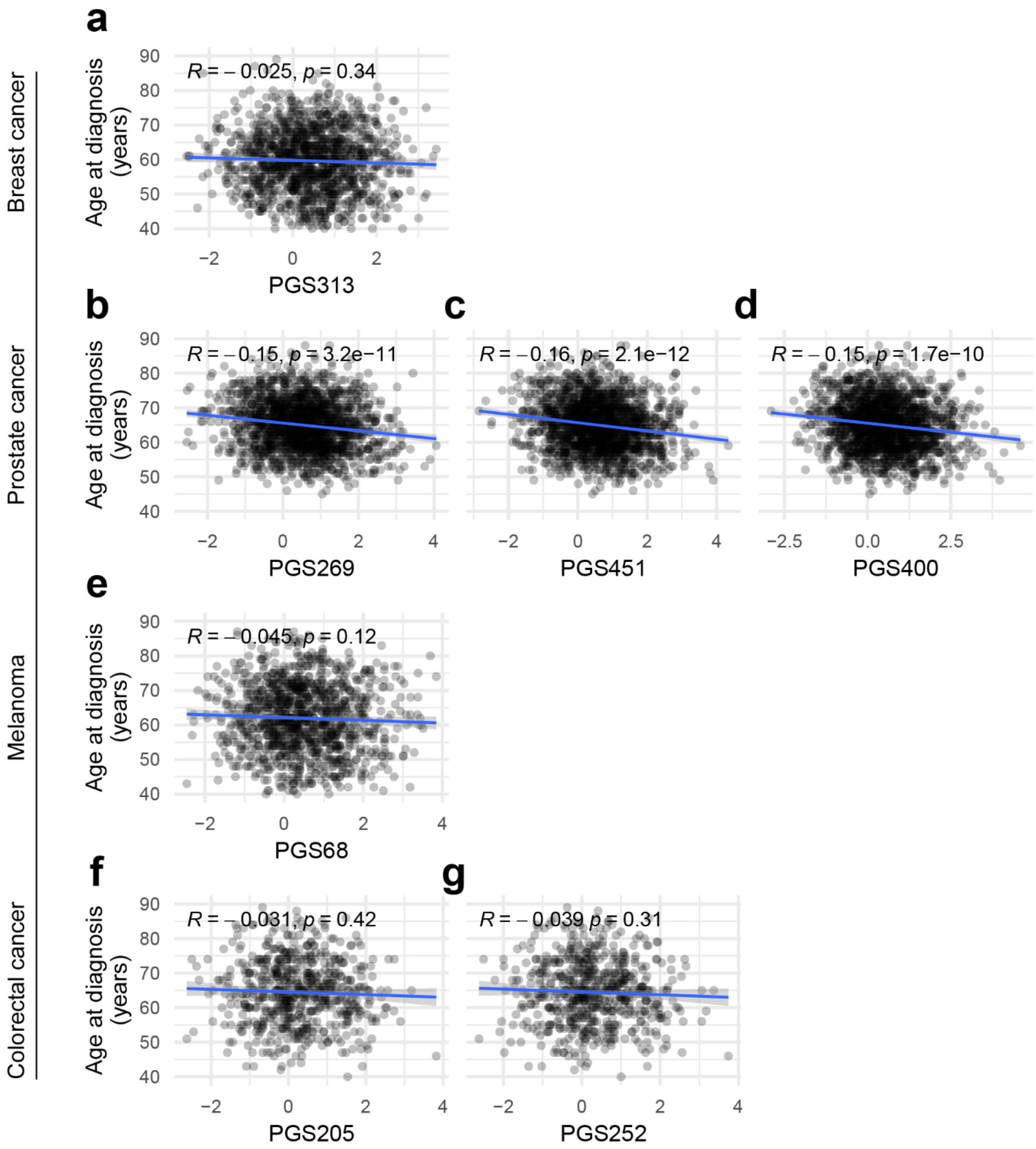

**Supplementary Figure 8. Association between polygenic scores and age at cancer diagnosis. (a–g)** Association between standardised polygenic scores (PGS) and age at cancer diagnosis among cases, assessed using Spearman correlation. Results are shown for breast cancer (**a**, PGS313), prostate cancer (**b–d**, PGS269, PGS451 and PGS400), melanoma (**e**, PGS68), and colorectal cancer (**f,g**, PGS205 and PGS252). Each point represents an individual, and the blue line indicates the fitted linear trend. Spearman correlation coefficients (*R*) and corresponding P values are shown in each panel. All analyses were restricted to unrelated European-ancestry individuals; for breast cancer and prostate cancer, analyses were further restricted to female and male participants only. Numbers of included participants with cancer diagnosis ("cases") are shown in Table 2.

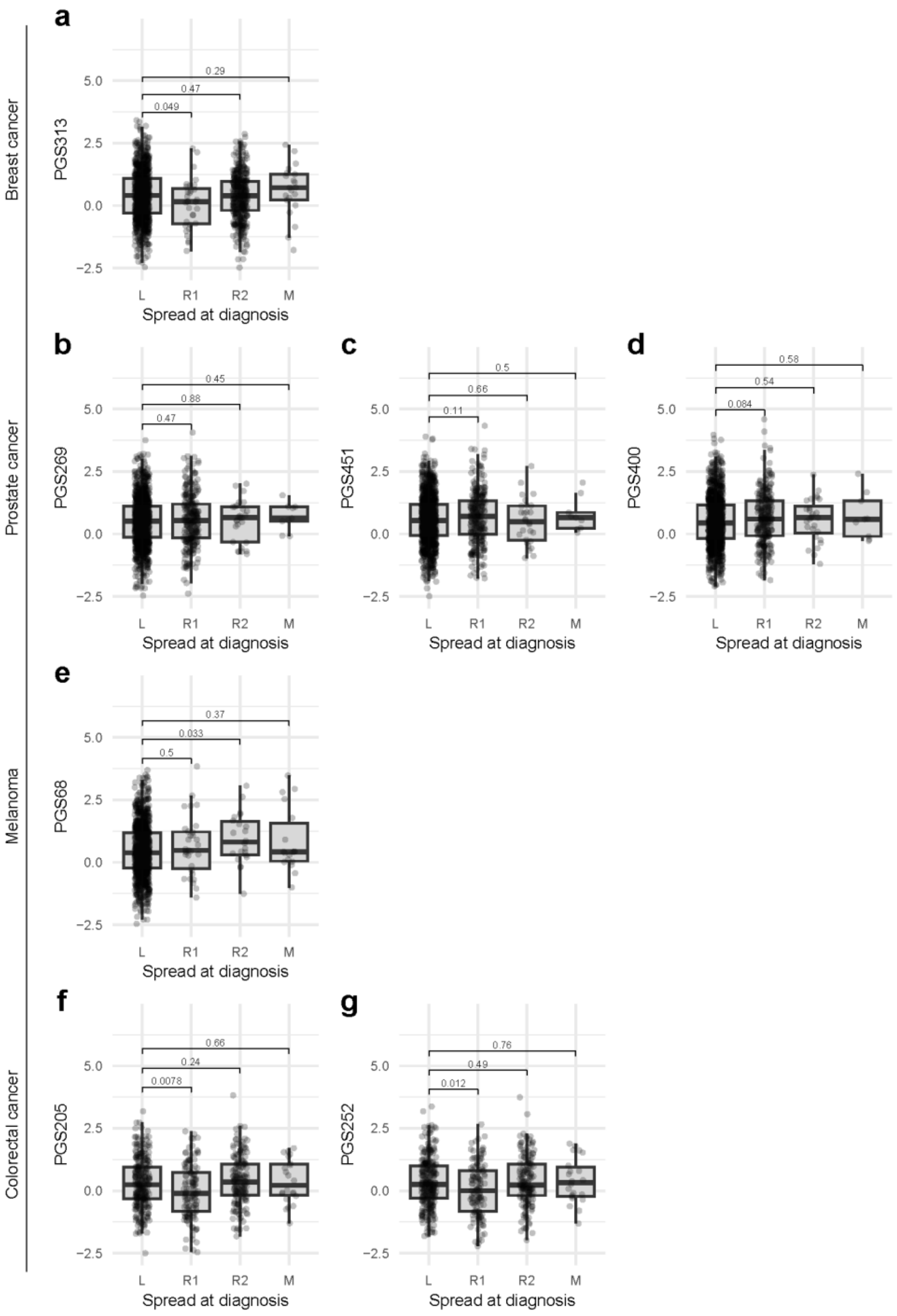

**Supplementary Figure 9. Association between polygenic scores and stage at cancer diagnosis. (a–g)** Distribution of standardised polygenic scores (PGS) among cases stratified by stage at diagnosis. Results are shown for breast cancer (**a**, PGS313), prostate cancer (**b–d**, PGS269, PGS451 and PGS400), melanoma (**e**, PGS68), and colorectal cancer (**f,g**, PGS205 and PGS252). Spread of cancer was categorised as localized (L), regional spread to adjacent organs (R1), regional spread to lymph nodes (R2), and distant metastases (M). Differences between groups were assessed using the Wilcoxon rank-sum test; *P* values for pairwise comparisons relative to the localized (L) stage are shown. All analyses were restricted to unrelated European-ancestry individuals; for breast cancer and prostate cancer, analyses were further restricted to female and male participants only. Of participants with cancer diagnosis (“cases”) as shown in Table 2, we excluded those with cancer registry record not including spread of disease at diagnosis (breast cancer: 48; prostate cancer: 437; melanoma: 55; colorectal cancer: 64).

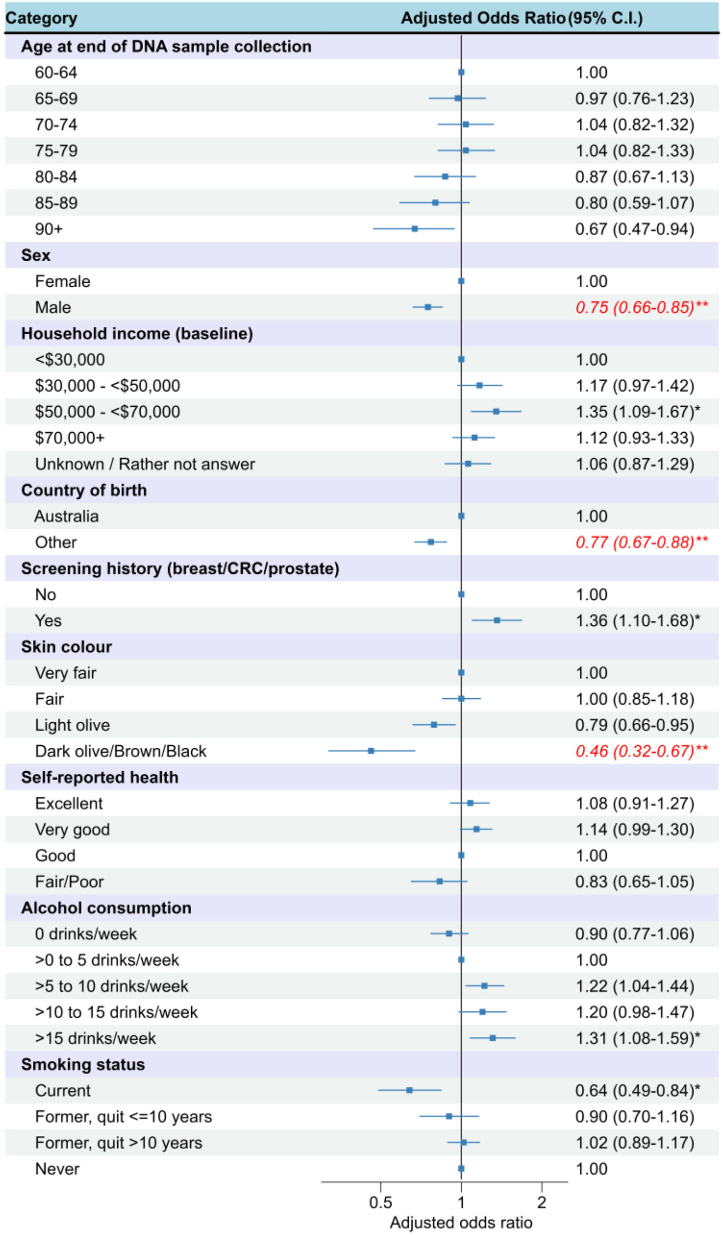

**Supplementary Figure 10. Associations with inclusion in final high-quality genomic dataset (n=6,827), among all individuals who consented to provide a DNA sample.** Regression analysis with simultaneous adjustment for all characteristics shown in the figure, based on step-wise forward selection (p<0.1). Characteristics not retained by step-wise forward selection (p>0.1) were health insurance status, area-level index of socioeconomic disadvantage, remoteness of place of residence, education level, study invitation group, body mass index, tannability of skin, hours spent outdoors, family history of six cancers, and personal history of heart disease/stroke, high blood pressure, diabetes, blood clots or depression/anxiety. This analysis was restricted to people who consented to provide a DNA sample (n=8,311). For this group, we investigated associations between having high-quality genomic data (n=6,827, prior to any exclusions based on relatedness or inferred genetic ancestry) and a range of sociodemographic and health characteristics. After consenting to provide a DNA sample, 370 people did not return a saliva sample (n=7,941 were returned), 533 people returned a DNA sample but genomic data could not be generated e.g. due to low DNA yield or not achieving sufficient coverage or not passing initial Gencove QC steps (n=7,408 had genetic data generated), and the genomic data did not pass all stringent quality checks for 581 people (n=6,827 people had high-quality genomic data).

** p<0.001; * p<0.01; C.I.: confidence interval; CRC: colorectal cancer.

#### 3. Supplementary Tables

**Supplementary Table 1. Participants' characteristics: variables and their categories from 45 and Up Study questionnaires and linked health data.**

| Characteristic / Category | [Source] Question / Response |
|---|---|
| Age at baseline | *[45 and Up Study baseline questionnaire] Derived from month and year of birth (supplied in 45 and Up Study dataset), number of years baseline date.* |
| Age at end of DNA sample collection | *[45 and Up Study baseline questionnaire] Derived from month and year of birth (supplied in 45 and Up Study dataset), number of years to 31 December 2023.* |
| Gender | *[45 and Up Study data] Gender: Female; Male* |
| Education | *[45 and Up Study baseline questionnaire] What is the highest qualification you have completed?* |
| University degree or higher | University degree or higher |
| Certificate/Diploma | Certificate/Diploma |
| Trade/Apprenticeship | Trade/Apprenticeship |
| High school | Higher school or leaving certificate (or equivalent) |
| Did not complete high school | School or intermediate certificate (or equivalent); No school certificate or other qualifications |
| Household income | *[45 and Up Study baseline questionnaire] What is your usual yearly HOUSEHOLD income before tax, from all sources? (please include benefits, pensions, superannuation, etc) [Tick box categories with income per year]* |
| <$30,000 | Less than $5,000; $5,000-$9,999; $10,000-$19,999; $20,000-$29,999 |
| $30,000 - <$50,000 | $30,000-$39,999; $40,000-$49,999 |
| $50,000 - <$70,000 | $50,000-$59,999; $60,000-$69,999 |
| $70,000+ | $70,000 or more |
| Unknown / Rather not answer | I would rather not answer this question; Missing values |
| Accessibility of place of residence | *[45 and Up Study baseline questionnaire] Remoteness area 2011 (ra_name_2011)* |
| Socioeconomic quintile | *[45 and Up Study baseline questionnaire] Deciles from the Index of Relative Socioeconomic Disadvantage (decile_rse_disadvantage) based on place of residence, aggregated to form quintiles* |
| Health insurance | *[45 and Up Study baseline questionnaire] Which of the following do you have? (excluding Medicare)* |
| Private health insurance | Private health insurance – with extras; Private health insurance – without extras |
| Department of Veterans' Affairs | Department of Veterans' Affairs white or gold card |
| Concession card | Health care concession card |
| None | None of these |
| Country of birth | *[45 and Up Study baseline questionnaire] In which country where you born* |
| Australia | Australia |
| Other | All other recorded values (any missing were set to missing) |
| Ancestry | *[45 and Up Study baseline questionnaire] What is your ancestry? (Please cross up to 2 boxes) Australian; English, Irish, Chinese; Italian; Greek; Scottish; German; Lebanese; Dutch; Maltese; Polish; Filipino; Indian; Croatian; Vietnamese; Other (please specify)* |
| Non-European category 1 | At least one of: Chinese; Filipino; Indian; Lebanese; Maltese; Vietnamese |
| Non-European category 2 | Category 1, plus "Other" (note: we did not have access to the free-text responses) |
| Family history of cancer | *[45 and Up Study baseline questionnaire] Have your mother, father, brother(s) or sister(s) ever had (blood relatives only):* |
| Breast cancer | Breast cancer |
| Colorectal cancer | Bowel cancer |
| Lung cancer | Lung cancer |
| Melanoma | Melanoma |
| Ovarian cancer | Ovarian cancer |
| Prostate cancer | Prostate cancer |
| Screening history | *[45 and Up Study baseline questionnaire]* |
| Breast cancer | Have you ever been for a breast screening mammogram? Yes/No |
| Colorectal cancer | Have you ever been screened for colorectal (bowel) cancer? Yes/No |
| Prostate cancer | Have you ever had a blood test ordered by your doctor to check for prostate disease (PSA test)? Yes/No |
| Skin colour | *[45 and Up Study baseline questionnaire] What best describes the colour of the skin on the inside of your upper arm, that is your skin colour without any tanning?* |
| Very fair | Very fair |

| | |
|---|---|
| Fair | Fair |
| Light olive | Light olive |
| Dark olive / Brown / Black | Dark olive; Brown; Black |
| Skin response to time in the sun (tannability) | *[45 and Up Study baseline questionnaire] What would happen if your skin was repeatedly exposed to bright sunlight during summer without any protection? Would it:* |
| Get very tanned | Get very tanned? |
| Get moderately tanned | Get moderately tanned? |
| Get mildly tanned | Get mildly or occasionally tanned? |
| Never tan, only freckle | Never tan, or only get freckled? |
| Outdoor hours | *[45 and Up Study baseline questionnaire] About how many hours a DAY would you usually spend outdoors on a weekday and on the weekend? [Weekday hours per day] [Weekend hours per day]* |
| *Quintiles 1-5* | Derived weekly exposure as 5x weekday hours plus 2x weekend hours, identified quintiles of weekly exposure, categorised each participant according to their weekly exposure value. |
| Self-reported health | *[45 and Up Study baseline questionnaire] In general, how would you rate your overall health?* |
| Excellent | Excellent |
| Very good | Very good |
| Good | Good |
| Fair / Poor | Fair; Poor |
| Personal history of diseases | *[45 and Up Study baseline questionnaire] Has a doctor EVER told you that you have:* |
| Heart disease / stroke | Heart disease; Stroke |
| High blood pressure | High blood pressure (female questionnaire specifies “when not pregnant”) |
| Diabetes | Diabetes |
| Depression / anxiety | Depression; Anxiety |
| Blood clots | Blood clot (thrombosis) |
| Body mass index | *[45 and Up Study baseline data] “bmi” values supplied in $kg/m^2$ based on weight and height* |
| Normal / Underweight | *Underweight (<18.5), normal (18.5-<25). Only ~1% of participants were underweight, so this category was combined with the normal category.* |
| Overweight | *25-<30* |
| Obese+ | *≥30* |
| Alcohol consumption | *[45 and Up Study baseline questionnaire] About how many alcoholic drinks do you have each week? One drink = a glass of wine, middy of beer or nip of spirits (put “0” if you do not drink, or have less than one drink each week)* |
| Categories (drinks/week) | 0; >0 to 5; >5 to 10; >10 to 15’>15; unknown |
| Smoking status | *[45 and Up Study baseline questionnaire]*<br>*1. Have you ever been a regular smoker?*<br>*2. If yes [to 1], are you a regular smoker now?*<br>*3. If no [to 2], how old were you when you stopped smoking regularly?* |
| Never | 1. No |
| Current | 1. Yes; 2. Yes |
| Former, quit ≤10 years | 1. Yes; 2. No; 3. Baseline age ≤ (Age stopped + 10) |
| Former, quit >10 years | 1. Yes; 2. No; 3. Baseline age > (Age stopped + 10) |
| Cancer type | *[NSW Cancer Registry] ICD-10 topography* |
| Breast | C50 |
| Colorectal | C18-C20 |
| Melanoma | C43 |
| Prostate | C61 |

**Supplementary Table 2. Demographic and health-related characteristics of 45 and Up Study participants invited, those who consented, those with high-quality genomic data, and unrelated individuals with high-quality data and inferred European genetic ancestry.**

| | Invited | | Consented | | High quality genomic data | | High quality data, unrelated, European ancestry | |
|---|---|---|---|---|---|---|---|---|
| **Characteristic** | **n** | **Column %** | **n** | **% of invited** | **n** | **% of invited** | **n** | **% of invited** |
| **Health insurance status (baseline)** | | | | | | | | |
| Private | 21,749 | 71% | 6,428 | 30% | 5,339 | 25% | 5,199 | 24% |
| Department of Veterans' Affairs | 251 | 1% | 43 | 17% | 31 | 12% | 31 | 12% |
| Healthcare card | 4,149 | 14% | 745 | 18% | 580 | 14% | 560 | 13% |
| None | 3,941 | 13% | 1,011 | 26% | 812 | 21% | 781 | 20% |
| Unknown | 451 | 1% | 84 | 19% | 65 | 14% | 60 | 13% |
| **Household income (baseline)** | | | | | | | | |
| <$30,000 | 7,554 | 25% | 1,526 | 20% | 1,202 | 16% | 1,164 | 15% |
| $30,000 -<$50,000 | 5,190 | 17% | 1,409 | 27% | 1,164 | 22% | 1,132 | 22% |
| $50,000 -<$70,000 | 3,609 | 12% | 1,145 | 32% | 972 | 27% | 946 | 26% |
| $70,000+ | 8,288 | 27% | 2,970 | 36% | 2,462 | 30% | 2,392 | 29% |
| Unknown/Rather not say | 5,900 | 19% | 1,261 | 21% | 1,027 | 17% | 997 | 17% |
| **Screening history** | | | | | | | | |
| Breast cancer | 13,576 | 44% | 3,714 | 27% | 3,122 | 23% | 3,020 | 22% |
| Colorectal cancer | 16,872 | 55% | 4,941 | 29% | 4,063 | 24% | 3,964 | 23% |
| Prostate cancer | 12,402 | 41% | 3,457 | 28% | 2,806 | 23% | 2,738 | 22% |
| Any of the above: Yes | 27,531 | 90% | 7,603 | 28% | 6,278 | 23% | 6,097 | 22% |
| Any of the above: No | 2,513 | 8% | 603 | 24% | 463 | 18% | 453 | 18% |
| Any of the above: Unknown | 497 | 2% | 105 | 21% | 86 | 17% | 81 | 16% |
| **Body mass index (kg/m$^2$)** | | | | | | | | |
| Normal/Underweight (<25) | 9,859 | 32% | 2,956 | 30% | 2,426 | 25% | 2,348 | 24% |
| Overweight (25-<30) | 12,206 | 40% | 3,334 | 27% | 2,745 | 22% | 2,668 | 22% |
| Obese+ (30+) | 6,416 | 21% | 1,539 | 24% | 1,267 | 20% | 1,233 | 19% |
| Unknown | 2,060 | 7% | 482 | 23% | 389 | 19% | 382 | 19% |
| **Alcohol consumption** | | | | | | | | |
| 0 drinks/week | 8,450 | 28% | 1,938 | 23% | 1,543 | 18% | 1,487 | 18% |
| >0, ≤5 drinks/week | 7,636 | 25% | 2,113 | 28% | 1,722 | 23% | 1,664 | 22% |
| >5, ≤ 10 drinks/week | 6,540 | 21% | 1,949 | 30% | 1,643 | 25% | 1,595 | 24% |
| >10, ≤ 15 drinks/week | 3,228 | 11% | 1,031 | 32% | 860 | 27% | 844 | 26% |
| >15 drinks/week | 4,294 | 14% | 1,211 | 28% | 1,012 | 24% | 999 | 23% |
| Unknown | 393 | 1% | 69 | 18% | 47 | 12% | 42 | 11% |
| **Smoking status** | | | | | | | | |
| Current | 1,557 | 5% | 337 | 22% | 251 | 16% | 243 | 16% |
| Former, quit ≤10 years | 1,819 | 6% | 451 | 25% | 365 | 20% | 360 | 20% |
| Former, quit >10 years | 8,523 | 28% | 2,357 | 28% | 1,955 | 23% | 1,905 | 22% |
| Never | 17,904 | 59% | 4,996 | 28% | 4,124 | 23% | 3,996 | 22% |
| Unknown | 738 | 2% | 170 | 23% | 132 | 18% | 127 | 17% |

| | | | | | | | | |
|---|---|---|---|---|---|---|---|---|
| **Skin colour** | | | | | | | | |
| Very fair | 4,846 | 16% | 1,384 | 29% | 1,155 | 24% | 1,134 | 23% |
| Fair | 17,146 | 56% | 4,816 | 28% | 4,007 | 23% | 3,936 | 23% |
| Light olive | 7,340 | 24% | 1,897 | 26% | 1,518 | 21% | 1,428 | 19% |
| Dark olive/Brown/Black | 889 | 3% | 155 | 17% | 103 | 12% | 94 | 11% |
| Unknown | 320 | 1% | 59 | 18% | 44 | 14% | 39 | 12% |
| **Tannability of skin** | | | | | | | | |
| Very | 7,888 | 26% | 2,199 | 28% | 1,786 | 23% | 1,703 | 22% |
| Moderate | 12,367 | 40% | 3,416 | 28% | 2,817 | 23% | 2,741 | 22% |
| Mild | 6,829 | 22% | 1,857 | 27% | 1,515 | 22% | 1,489 | 22% |
| Never | 2,892 | 9% | 723 | 25% | 611 | 21% | 605 | 21% |
| Unknown | 565 | 2% | 116 | 21% | 98 | 17% | 93 | 16% |
| **Outdoor hours (quintile)** | | | | | | | | |
| Quintile 1: Highest | 6,119 | 20% | 1,485 | 24% | 1,183 | 19% | 1,153 | 19% |
| Quintile 2 | 6,075 | 20% | 1,624 | 27% | 1,314 | 22% | 1,271 | 21% |
| Quintile 3 | 5,428 | 18% | 1,516 | 28% | 1,257 | 23% | 1,225 | 23% |
| Quintile 4 | 5,404 | 18% | 1,654 | 31% | 1,390 | 26% | 1,351 | 25% |
| Quintile 5: Lowest | 5,644 | 18% | 1,675 | 30% | 1,395 | 25% | 1,354 | 24% |
| Unknown | 1,871 | 6% | 357 | 19% | 288 | 15% | 277 | 15% |
| **Cancer type, including randomly selected cohort*** | | | | | | | | |
| Breast cancer | 5,981 | 20% | 1,719 | 29% | 1,439 | 24% | 1,400 | 23% |
| Colorectal cancer | 3,153 | 10% | 756 | 24% | 617 | 20% | 597 | 19% |
| Melanoma | 4,844 | 16% | 1,373 | 28% | 1,149 | 24% | 1,124 | 23% |
| Prostate cancer | 7,896 | 26% | 2,259 | 29% | 1,828 | 23% | 1,788 | 23% |
| Any of the above | 21,874 | 72% | 6,107 | 28% | 5,033 | 23% | 4,909 | 22% |

** Each individual assigned to group based on the first cancer diagnosis*.

**Supplementary Table 3. Sensitivity analysis comparing PGS performance using all variants present in the imputed VCF and the post-QC variant set (n=6,631 unrelated individuals with inferred European genetic ancestry and high-quality genomic data).** For each PGS, the number of variants in the original score, the number present in the imputed VCF, and the number retained after quality control (post-QC) are shown. Discriminative performance is reported as the area under the receiver operating characteristic curve (AUC) with 95% confidence intervals using (i) all PGS variants present in the imputed VCF and (ii) post-QC variants only.

| PGS ID | Cancer type (n total; n cases) | Variants (original; in VCF; post-QC) | AUC (95% CI), using variants in VCF | AUC (95% CI), using post-QC variants |
|---|---|---|---|---|
| PGS313 | Breast (2,487; 1,431) | 313; 260; 218 | 0.629 (95% CI 0.608-0.651) | 0.618 (95% CI 0.596-0.64) |
| PGS269 | Prostate (2,642; 1,879) | 269; 236; 196 | 0.669 (95% CI 0.647-0.692) | 0.658 (95% CI 0.635-0.681) |
| PGS451 | Prostate (2,642; 1,879) | 451; 390; 337 | 0.689 (95% CI 0.667-0.711) | 0.68 (95% CI 0.658-0.703) |
| PGS400 | Prostate (2,642; 1,879) | 400; 348; 302 | 0.665 (95% CI 0.643-0.688) | 0.663 (95% CI 0.64-0.685) |
| PGS68 | Melanoma (3,150; 1,218) | 68; 60; 51 | 0.644 (95% CI 0.625-0.664) | 0.638 (95% CI 0.618-0.658) |
| PGS205 | CRC (2,633; 661) | 205; 166; 127 | 0.59 (95% CI 0.566-0.615) | 0.569 (95% CI 0.544-0.595) |
| PGS252 (PGS205 plus 47 fine-mapped SNPs) | CRC (2,633; 661) | 252; 205; 158 | 0.597 (95% CI 0.573-0.622) | 0.575 (95% CI 0.55-0.6) |
